# Early phase of the COVID-19 outbreak in Hungary and post-lockdown scenarios

**DOI:** 10.1101/2020.06.02.20119313

**Authors:** Gergely Röst, Ferenc A. Bartha, Norbert Bogya, Péter Boldog, Attila Dénes, Tamás Ferenci, Krisztina J. Horváth, Attila Juhász, Csilla Nagy, Tamás Tekeli, Zsolt Vizi, Beatrix Oroszi

## Abstract

COVID-19 epidemic has been suppressed in Hungary due to timely non-pharmaceutical interventions, prompting a huge reduction in the number of contacts and transmission of the virus. This strategy was effective in preventing epidemic growth and reducing the incidence of COVID-19 to low levels. In this report, we present the first epidemiological and statistical analysis of the early phase of the COVID-19 outbreak in Hungary. Then, we establish an age-structured compartmental model to explore alternative post-lockdown scenarios. We incorporate various factors, such as age-specific measures, seasonal effects, and spatial heterogeneity to project the possible peak size and disease burden of a COVID-19 epidemic wave after the current measures are relaxed.

## 1. Introduction

A cluster of pneumonia cases of unknown origin was detected in Wuhan city, the capital of Hubei Province, China, with a population of 11 million in December 2019. On December 31, 2019 China alerted the World Health Organization (WHO) China Country Office [1]. Epidemiological investigation to find common exposure implicated that many early cases of pneumonia of unknown aetiology were associated with the Huanan Seafood Wholesale Market that was subsequently closed for environmental sanitation and disinfection. As part of the epidemiological investigation, active case finding was initiated [2,3]. On January 7, 2020 the causative pathogen of the pneumonia outbreak was identified as a novel coronavirus (named SARS CoV-2), and on January 12, China shared the genetic sequence with the public.

It is well known, that some coronaviruses are circulating in animals and others in humans. It was recognized that SARS-CoV-2 apparently succeeded last year in making its transition from animals to humans in China. Similar successful mutations in coronavirus have already happened several times in the 21st century. So far, we have seen two other major highly pathogenic zoonotic outbreaks of coronaviruses. The first was Severe Acute Respiratory Syndrome Coronavirus (SARS-CoV) in 2002 that infected over 8,000 people within a short time period and had a case fatality rate (CFR) close to 10% [4]. The second, Middle Eastern Respiratory Syndrome (MERS) Coronavirus in 2012, was more difficult to transmit, but had a substantially higher CFR over 30% [5].

COVID-19 is less fatal than any of these predecessors, but, on the other hand, this new coronavirus has a great potential for transmission. When the first devastating outbreak in Wuhan happened, the world watched the unprecedented scale of quarantine restrictions in China. However, as more and more cases were identified outside China, it was clear that it is not feasible to suppress the epidemic at the source, so countries started to strengthen their surveillance to be able to quickly identify potential cases. In this early phase, case definitions, protocols for diagnosis and treatment, management of close contacts were being developed, and effort was made to increase alert to ascertain cases and determine if it was of imported origin or resulted from a within country transmission.

Coronaviruses affect mainly the respiratory tract. Based on Chinese data, about 80% of laboratory confirmed patients reported until February 20,2020 had mild to moderate disease including non-pneumonia and pneumonia cases. Mild symptoms are like the common cold, including sudden onset feverishness, fatigue, runny nose and cough. 13.8% of cases were severe and 6.1% were critical (defined as respiratory failure requiring mechanical ventilation, shock or other organ failure that requires intensive care). The WHO-China Joint Mission reports that 75% of initially asymptomatic cases progressed to clinical disease in China, hence, true asymptomatic infection was estimated to be rare (1–3%) [6]. Severe illness and death were more likely to affect people aged over 60 years or those with underlying conditions. Disease in children appeared to be relatively rare and mild [3].

The first three cases in Europe, with a recent stay in Wuhan, were confirmed on January 24, 2020 in France (where, later in April, COVID-19 was retrospectively confirmed for a patient hospitalised in late December 2019) [7,8]. Within a few days, two Chinese tourists were confirmed with COVID-19 in Italy on January 31.

The first epidemic in Europe started in the Lombardy region of Italy with the first detection on February 20, 2020. During the last week of the same month, the number of cases increased rapidly within a few days, half of which required hospitalisation. The rapid intensification of regional surveillance, by tracing and testing all known patients’ contacts, revealed that the epidemic had already spread in the southern part of the region and found the earliest transmission in January 2020 [9]. On March 8, the Italian government introduced containment measures in Lombardy and neighbouring provinces, then, on March 11, in the whole country.

Those measures were successful in preventing the rise of the epidemic in central and southern Italy, while in the northern regions (Lombardy, Emilia-Romagna and Veneto) the epidemic had already reached high levels by mid-March [10].

The WHO Director-General declared the COVID-19 outbreak a Public Health Emergency of International Concern under International Health Regulations (2005) on January 30, 2020 [11] and then, a pandemic on March 11,2020 [12]. By that time more than 118,000 cases were detected in 114 countries and 4,291 people died in COVID-19.

Globally, it took about 3 months to reach 1 million confirmed cases, but then, the cumulative incidence doubled just in two weeks and by April 28, the reported number of COVID-19 cases reached 3 million. According to the WHO, by May 10, 2020, there were over 4 million confirmed cases of COVID-19 and over 278,000 fatalities worldwide. Both Europe and the Americas had over 1.7 million cases and over 150,000 and 100,000 fatalities, respectively [1,13].

From March 2020, most European states implemented strict physical distancing measures such as school and workplace closures, cancellation of public events, restrictions of gatherings, and stay-at-home requirements as a response to the pandemic, aiming to reduce transmission of SARS-CoV-2. These measures reflect an extraordinary effort to suppress, or at least to slow down COVID-19, and they proved to be effective.

By May 2020, some European countries show signs of improvement as their number of the daily new cases has begun to decline: in Italy from the end of March, in Austria from the second week of April, and in mid May, similar decline can be observed in the UK, France, and Spain. As a consequence of reducing the transmission of SARS-COV-2, from the second half of April several countries (e.g. Austria, Denmark, and Germany) have started to gradually raise their mitigation measures by, for example, re-opening primary schools, restaurants, and shops.

Mathematical models have been developed to better understand the global spread [14] and the transmission dynamics of COVID-19 for many countries, including Australia [15], France [16], Germany [17], UK [18], USA [19]. Such models have been used to project the evolution of the outbreak and to estimate the impact of control measures on reducing disease burden. Here, we provide the first epidemiological and statistical analysis of the early phase of the COVID-19 outbreak in Hungary, then, we employ an age stratified compartmental model to explore possible future scenarios.

## 2. Methods

### 2.1. Epidemiological report

During the last week of January 2020, the National Public Health Center (hereinafter NPHC) issued the surveillance protocol of COVID-19 cases that included the case definitions of COVID-19 cases. Till mid May the protocol was updated four times as the relevant WHO and/or ECDC guidance was revised. According to the Hungarian protocol, suspected cases were to be reported by physicians to the local health authority and NPHC, followed by compulsory testing, 14-day isolation and monitoring, case investigation, and contact tracing. A person with laboratory confirmation (detection of SARS-CoV-2 by polymerase chain reaction), irrespective of clinical signs and symptoms, was considered a confirmed COVID-19 case.

The first confirmed COVID-19 cases were reported during the first week of March 2020 through the Hungarian Notifiable Disease Surveillance System operated by NPHC. The first case, a foreign university student studying and residing in Hungary, was reported on March 4, 2020. By May 10, 2020, the cumulative number of reported confirmed COVID-19 cases were 3284 (33.1 cases per 100,000 population), including 421 deaths (crude CFR 12.8%), see Figure 1 for the daily reported numbers. Out of the 3284 cases, 47.0 % (1542 cases) occurred in the 65+ age group, 29.1% (957 cases) in the 20–49 age group, 21.9% (718 cases) in the 50-64 age group and 2.0% (67 cases) among people under 20-year. Age specific morbidity was highest in the 80+ age group (163.3 cases per 100000 population) and more than twice of the overall in the 70–79 age group (69.5 cases per 100,000 population).

**Figure 1.**
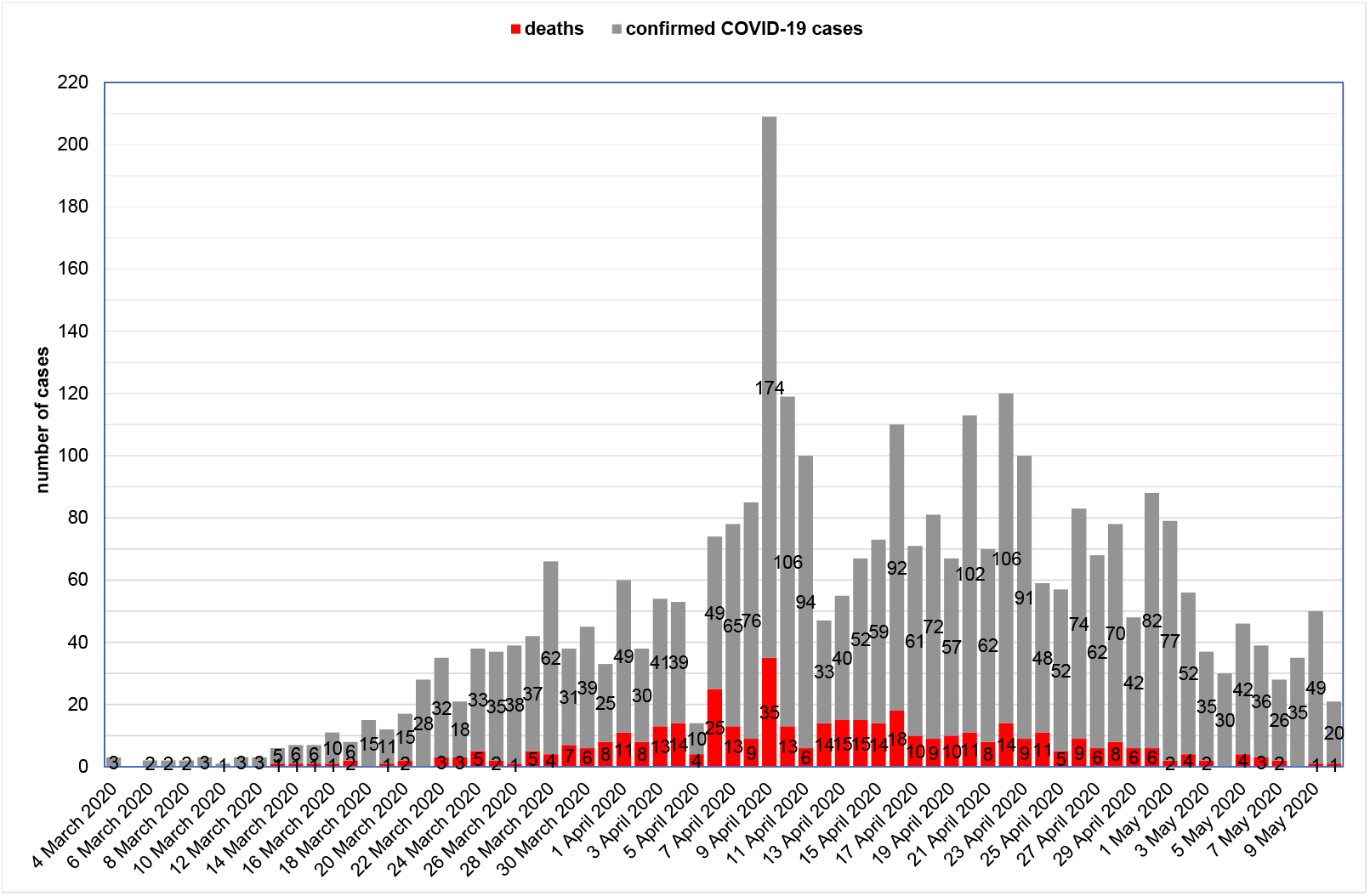
Epidemic curve of confirmed COVID-19 cases in Hungary by date of confirmation (reported until May 10, 2020)

**Figure 2.**
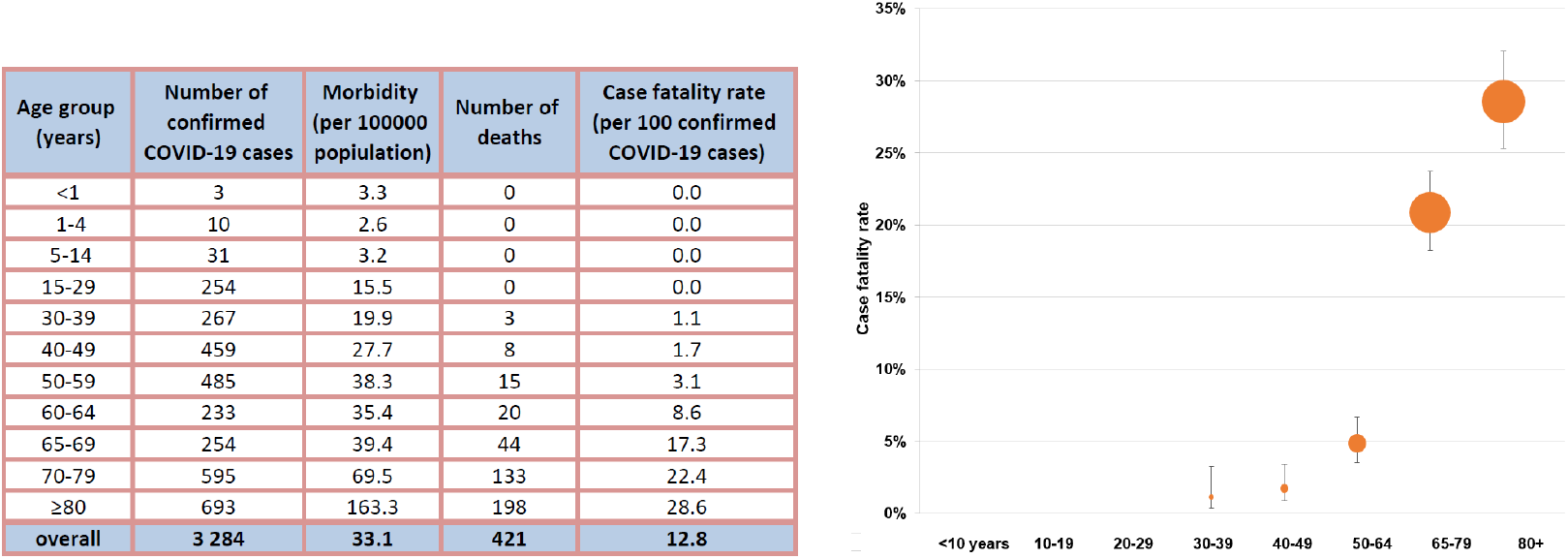
Crude case fatality rate of confirmed COVID-19 cases in Hungary by age groups (reported until May 10)

Out of 421 deaths, 89.1% (375 deaths) belonged to the 65+ age group. The highest crude CFR was observed in the 80+ age group (28.6%), followed by the 70–79 age group (22.4%) and the 65–69 age group (17.3%). No deaths were reported under 33 years of age.

Out of the 3284 cases, 58.0% (1906 cases) were female and 41.9% (1376 cases) male (gender not reported for two cases). The morbidity among women was higher (37.1 vs. 28.8 cases per 100,000 population), so men were 0.8 (95% CI 0.72–0.83) less likely to become ill. However, men aged 33 years and older had a 1.2 (95% CI 0.96–1.41) higher risk to die than women aged 33 years and older (7.1 cases vs. 6.0 cases per 100,000 population). Out of 3284 cases, at the stage of data consolidation as of May 10, 2020, we have information about the symptoms of 63.2% (2076 cases). Out of 2076 cases, 29.5% (613 cases) had no symptoms, 52.3% (1086 cases) had mild symptoms, 18.2% (377 cases) had severe disease (including 149 cases required intensive care and/or ventilation).

Most of the cases were reported from the central part of Hungary, from the capital (1587 cases), Pest county (438 cases) and Fejér county (333 cases). The morbidity (per 100,000 population) was the highest in Budapest (94.7), followed by Fejér county (80.1) in central Hungary and Zala county (60.8) in western Hungary.

Between March 4 and April 8, there was a slow increasing trend in the daily reported number of new cases (varying between 1 and 85 cases) peaked on April 9 (209 cases). After a period from April 10 till April 22 with fluctuating number of daily reported new cases (between 47 and 120 cases), starting from April 23 the number of daily new cases decreased from 100 to 21 cases per day. The 14-day cumulative number of new cases steadily increased until its peak on April 22 (13.1 per 100,000 population) before it started to decrease and was just over 10.0 cases per 100,000 population between April 11 and May 3, 2020.

The epidemic curve reflects a propagated source epidemic especially when we consider only those cases that cannot be connected to outbreaks in closed communities (like long-term care facilities or hospitals) or to health care associated infections. Out of 3284 cases, 31.4% (1031 cases) were associated with health care and/or outbreaks in hospitals, contributing to the daily reported new cases since mid-March. Health care workers had 10.0 times (95% CI 9.02-10.99) higher risk to become a confirmed COVID-19 case in comparison to the general population (288.8 cases vs. 29.0 cases per 100,000 population). Out of 3284 cases, 27.8% (913 cases) were reported from long-term care facilities (nursing homes and other closed communities like homeless shelters) contributing to the daily reported new cases since early April. At the peak of the epidemic curve, 62.2% (130 cases) of cases on April 9 were reported from the same retirement and assisted living facility.

### 2.2. Statistical analysis

Statistical analysis of the available surveillance data – such as case counts and death counts – is an indispensable tool during outbreak response. It can reveal the current situation objectively, shed light on the effect of past interventions, uncover non-obvious aspects of the outbreak and produce forecasts. It can also provide important input information to disease dynamics models.

This section summarizes the statistical analyses carried out during the early phase of the COVID-19 outbreak in Hungary, pointing out the most important tasks and results.

All methods were implemented under the R statistical environment version 4.0.0 [20] using packages ggplot2 version 3.3.0 [21] for visualization, data.table version 1.12.8 [22] for data manipulation and shiny version 1.4.0.2 [23] for creating an interactive dashboard to carry out epidemiological analyses online (available in Hungarian [24]).

The full source code of of this dashboard and every analysis presented in this section is available at [25].

#### 2.2.1. Temporal variation of the effective reproduction number

Effective reproduction number (*R_t_*), the average number of secondary cases per primary case for those primary cases who turn infectious on day *t*, was tracked real-time based on the daily number of reported new cases using the methods of Cori *et al*. [26] and that of Wallinga and Teunis [27], *inter alia*. In brief, the method of Cori *et al*. is based on calculating the ratio of the actual number of infections on a day to the total infectiousness of all past cases on that day. Thus, it measures *R_t_* by assuming that infected individuals will infect in the future as if conditions remain unchanged. The method of Wallinga and Teunis uses a likelihood-based inference on the possible infection networks underlying the epidemic curve.

The fundamental difference is that method of Cori *et al*. solely uses past information (“backward looking approach”), the reason for the result being sometimes called instantaneous reproduction number, while the Wallinga–Teunis method more closely corresponds to the concept of the usual definition of effective reproduction number, however, it requires future information in exchange (“forward looking approach”). For a discussion on the relative merits of these two approaches see [28,29].

Figure 3. shows the results. The reproduction number showed a steady decline - apart from an outlying effect in early April - and became close to, or even below 1 by mid-April, and remained at that level since then. This conclusion is robust to the chosen methodology.

**Figure 3.**
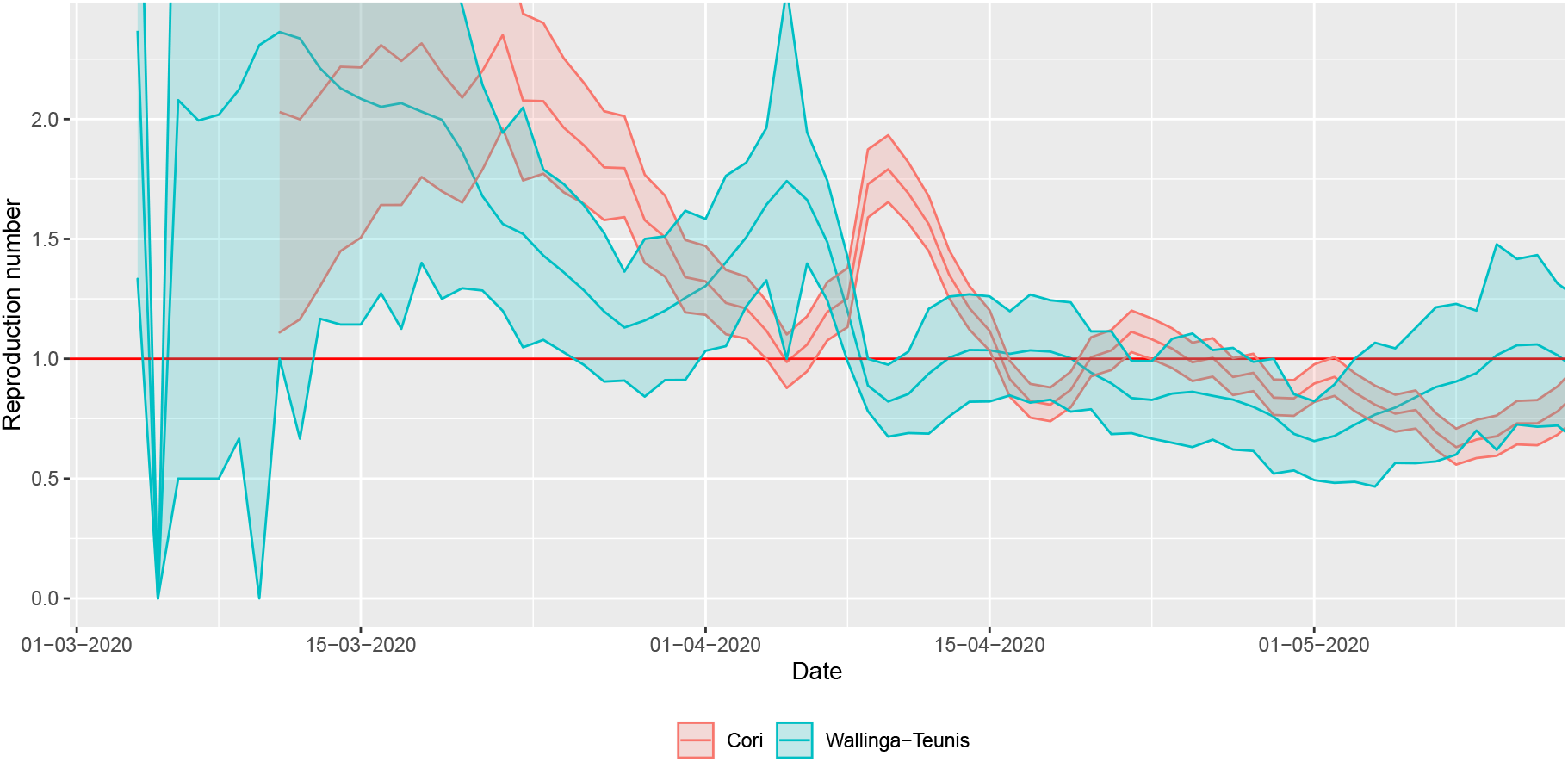
Real-time estimation of the reproduction number during the early phase of the COVID-19 outbreak in Hungary using two different methods (shaded area depicts 95% confidence interval).

Both methods require – in addition to incidence data – information on the serial interval. Depending on the used dataset, different estimations of the serial interval have been published: a mean of 3.96 days was found in [30], and 6.6 days in [31]. Here we assume an intermediate value following [32], where the mean and standard deviation (SD) of the serial interval were estimated at 4.7 days (95% CrI: 3.7, 6.0) and 2.9 days (95% CrI: 1.9,4.9), respectively. (The serial interval is assumed to follow gamma distribution.) They also concluded that the serial interval of COVID-19 is close to or shorter than its median incubation period, which is coherent with our choice of parameters in the transmission dynamics model.

The estimation was carried out using `R packages R0` version 1.2-6 [33,34] and `EpiEstim` version 2.2-2 [26,35].

#### 2.2.2. Adjusted case fatality ratio

Case fatality rate (CFR) is defined as the (conditional) probability of death from a disease for those contracting the disease (for diseases where asymptomatic state also exists, infection fatality rate, IFR, is defined analogously) and is estimated as the ratio of cumulative deaths and cumulative cases. This definition, i.e.,

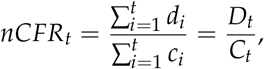

where *c_t_* and *d_t_* are the daily, *C_t_* and *D_t_* are cumulative number of cases and deaths, respectively, on day *t* is however biased when used during the epidemic (thus the name naive CFR or nCFR). The reason for this is that a proportion of cases counted in the denominator will die (in the future), thus they should have been counted in the numerator as well, but as they’re not, the ratio underestimates the true value [36,37].

Fortunately, it is relatively easy to correct for this bias using information on the distribution of the diagnosis-to-death time [38,39]. Denoting by *f_i_* the (conditional) probability that death happens on day *i* after the onset for those who die, the likelihood that the cumulative number of deaths on day *t* is *D_t_* is

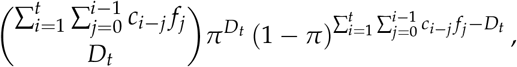

where *π* denotes the true value of the CFR. This observation allows both maximum likelihood and Bayesian estimation for using the observed series of *c_i_* and *d_i_*, the latter of which was employed in the present study, using a Beta(1,1) (i.e., uniform) prior. It was assumed that diagnosis-to-death time follows lognormal distribution with a mean of 13 days and a standard deviation of 12.7 days, as found by Linton *et al*. [40].

Results are shown on Figure 4. Note that – as the outbreak is coming to its end – the naive method converges to the final value that was readily well estimated almost a month earlier by the corrected technique. This case fatality rate is about 16%.

**Figure 4.**
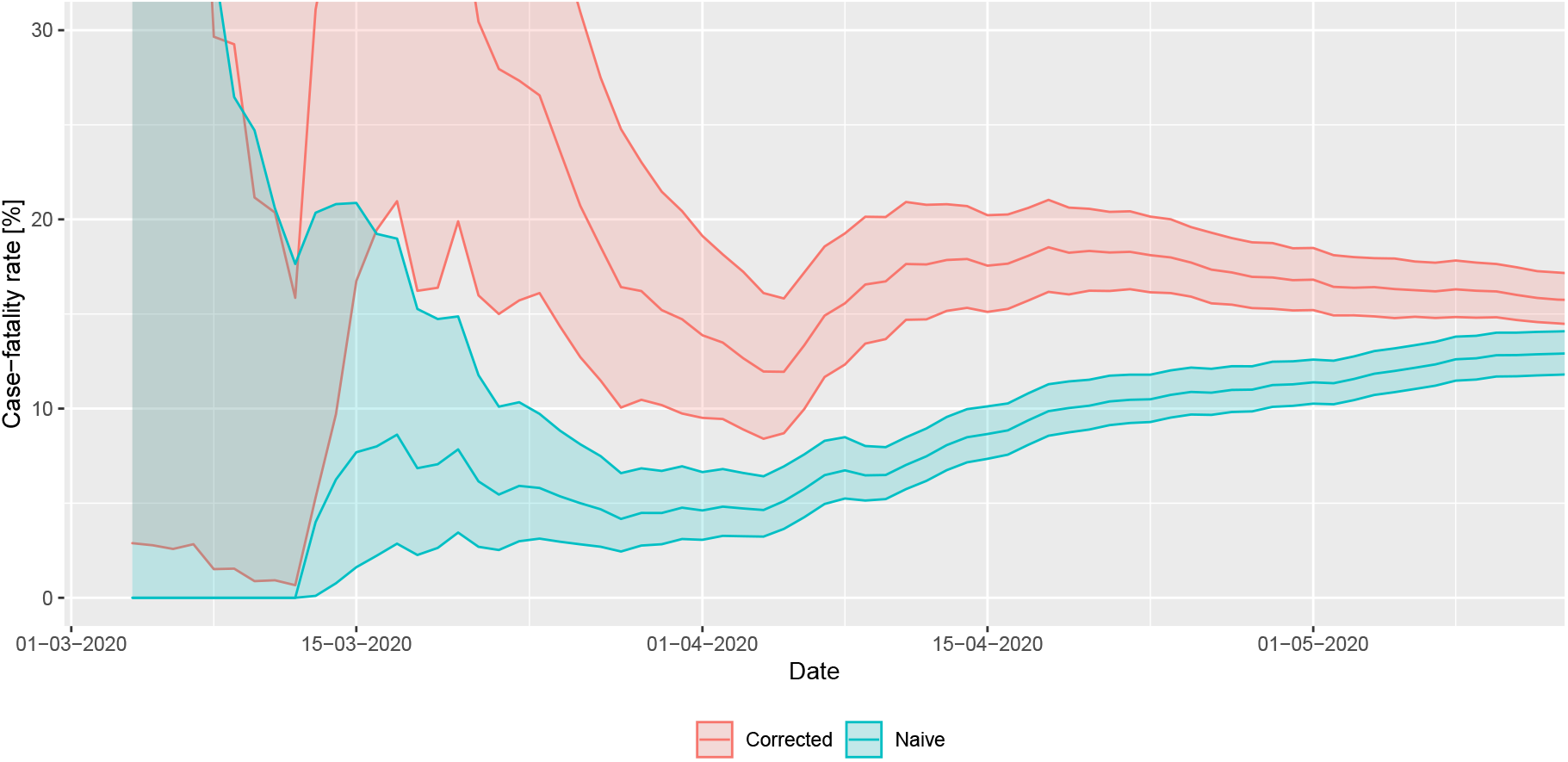
Real-time estimation of case fatality rate during the early phase of the COVID-19 outbreak in Hungary (shaded area depicts 95% confidence interval).

The Bayesian estimation was manually coded using the `R package rstan` version 2.19.3 [41]. Markov chain Monte Carlo approach was used to carry out the estimation with No-U-Turn sampler, using 4 chains, 1000 warmup iterations and 2000 iterations for each chain.

#### 2.2.3. Estimation of ascertainment rate

The 16% case fatality rate mentioned in the previous subsection is still not the true value, as there is another source of bias, but this time leading to overestimation: the underdetection of cases. This is a substantial issue now as a – precisely not yet known, but epidemiologically significant – fraction of the COVID-19 cases are asymptomatic or mild symptomatic. Since in many countries testing was extended to contacts (and in a few instances, even random sampling was carried out), the confirmed cases include some asymptomatic cases as well.

However, the value of the estimated (corrected) infection fatality rate can also be used to estimate the ascertainment rate: by assuming that the IFR in reality takes a benchmark value (one derived from large-sample, well-designed studies accounting for underdetection or sero-epidemiological surveys) and – crucially – assuming that the difference of the actual estimated IFR from that value is purely due to underdetection, the ascertainment rate can be obtained by simply dividing the assumed true value of the IFR with the actual estimated CFR [42]. Note that this might be a strong assumption, as it rules out that there is a real difference in the country’s IFR from the benchmark value, in particular, it rules out different virulence of the pathogen, different age- and comorbidity-composition in the country and different effect of the healthcare system on survival.

Various IFR estimations have been published, for example 0.66% for China [43], 0.9% for UK [18]. Recent serological studies found IFR values spanning from 0.36% in a German town [44], to 1.19% in Milan [45]. Here we explore a reasonable range of IFRs from 0.3% to 1.2%, and the results are shown in Table 1. Note that earlier estimates based on [43] and [18] are consistent with the preliminary results of a large-scale Hungarian sero-epidemiological study [46].

**Table 1.** Underdetection (ratio of all infections to reported cases) and corrected number of cumulative cases based on the estimated underdetection.

| IFR | 0.3% | 0.6% | 0.9% | 1.2% |
| --- | --- | --- | --- | --- |
| Underdetection (true/reported) | 54.0 | 27.0 | 18.0 | 13.5 |
| Corrected cumulative number of infections by May 10 | 177,242 | 88,621 | 59,081 | 44,310 |

### 2.3. Description of the transmission model

We establish a compartmental population model, adjusted to the specific characteristics of COVID-19. Several studies [15-17,19,47] have proposed similar models for the transmission dynamics of COVID-19. We consider the following compartments. We denote by *S* the susceptibles, i.e. those who can be infected by the disease. Latent (*L*) are those who have already contracted the disease but do not show symptoms and are not infectious yet. In accordance with studies indicating that viral shedding peaks before the onset of symptoms [48], in our model, we have introduced the presymptomatic infected compartment *I***_p_** for those who do not have symptoms, but who already are capable to transmit the disease to susceptibles. We divided the latent period into two compartments *L***_1_** and *L***_2_**, thus together with *I***_p_**, the incubation period follows a hypoexponential distribution, having a shape matching empirical observations [49]. Since a large fraction of infected show only mild or no symptoms, after the incubation period, we differentiate these individuals from those with symptoms. We assume a gamma-distributed infectious period with Erlang parameter *m* = 3, following [50], hence, we have three classes for both asymptomatic and symptomatic infectious individuals (*I***_a,1_**, *I***_a,2_**, *I***_a,3_** and *I***_s,1_**, *I***_s,2_**, *I***_s,3_**, respectively). Individuals from the *I***_a,3_** compartment will all recover and hence proceed to the recovered class *R*. Immunity is assumed for those who have recovered from the disease, at least for the time scale of this modelling. Individuals from *I***_s,3_** may either recover without requiring hospital treatment (and thus move to *R*) or become hospitalized. It is of crucial importance to project the number of hospital beds and ICU beds needed, thus in the model we further differentiate symptomatically infected individuals who need hospital care and critical care, denoted by *I***_h_** and *I***_c_**, respectively. We operate with the assumption that the healthcare system will not be overwhelmed, and thus disease-induced death is only considered from critical care, hence, individuals from *I***_h_** will proceed to *R* after recovery. Those from *I***_c_** with fatal outcome transit to the *D* compartment. Those who are out of ICU and on the path to recovery are collected into the *I***_cr_**, from where they eventually recover and move to the *R* class.

To take into account the different characteristics of the disease in various age groups, we stratified the Hungarian population into seven groups, corresponding to the eligible age groups in the Hungarian online questionnaire for the assessment of changes in the number of contacts following the lockdown [51]. The compartments listed above corresponding to the different age groups are denoted by an upper index *i* ∈ 1, ..., 7. Accordingly, all of our parameters can be calibrated age-specifically.

The transmission rates from age group *k* to age group *i* are denoted by 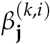, with **j** ∈{**p**, **a**, **s**}, where the three subscripts **p**, **a**, **s** stand for presymptomatic, asymptomatic, and symptomatic infected, respectively. The parameters described in the following all have an upper index *i* which stands for the corresponding age group. A fraction *p^i^* of exposed people will not show symptoms during his/her infection, while (1 − *p^i^*) will develop symptoms. The average length of the incubation period is 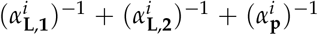 days, with the transition rates 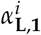, 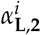, 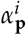, respectively. Similarly, the average infectious period of asymptomatic and symptomatic infected individuals are 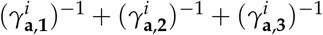, and 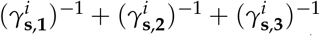, with the corresponding transition rates, respectively. A fraction *h^i^* of the infectious compartment 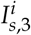 will be hospitalized, the remaining fraction 1 − *h^i^* will recover without hospital care. Out of those who need hospitalization, a fraction *ξ^i^* need intensive care. For the hospitalized classes 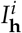, 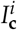, 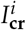, the average time spent in these compartments is given as 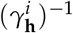, 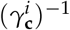 and 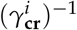, respectively. A fraction *µ^i^* of those leaving the 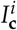 compartment will die due to the disease, while the remaining fraction will proceed to the 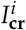class.

The transmission dynamics of our model for one age group is illustrated in Figure 5, while the governing system of differential equations (A1) of our model can be found in Appendix A. The model parameters with references are detailed in Appendix B. A further important component of our model is the contact matrix, describing social mixing between the age groups, which can be found in Appendix C. The elements of the contact matrix are included via the different transmission terms 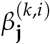. Reproduction numbers are calculated using the next generation matrix method in Appendix D.

**Figure 5.**
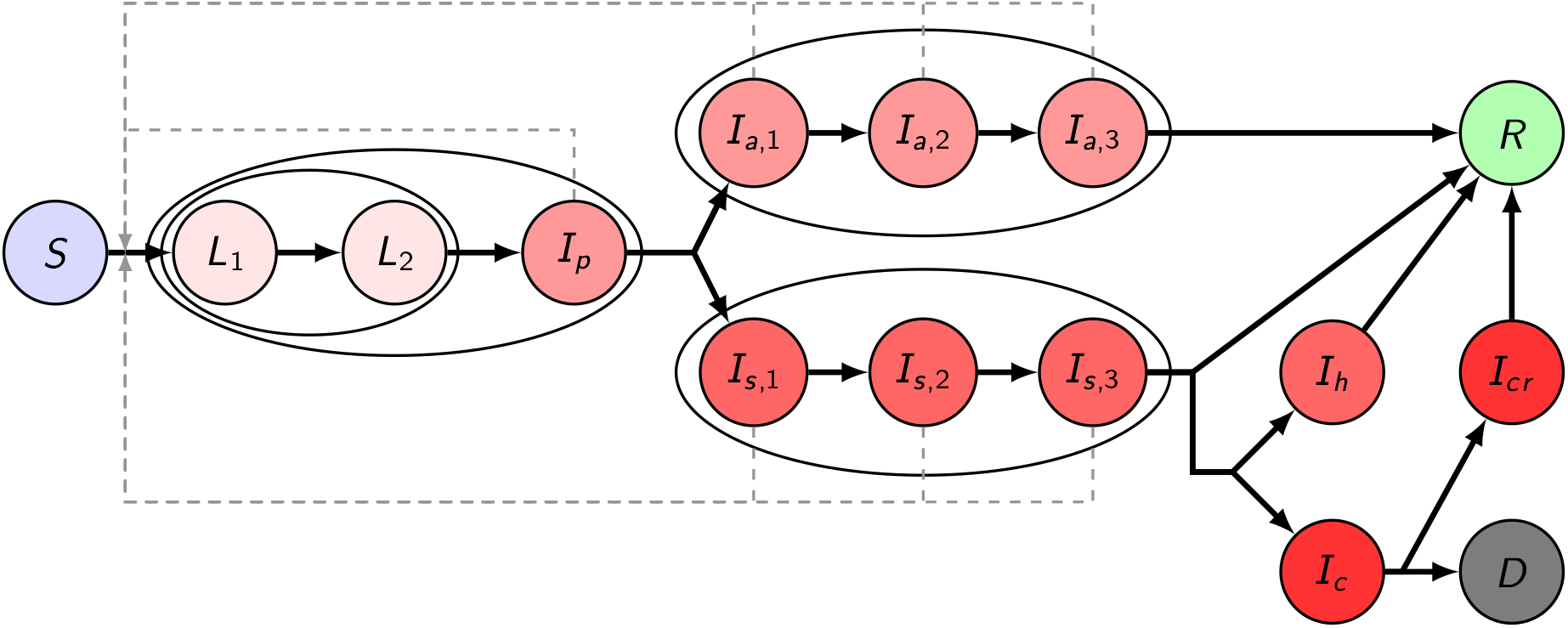
Transmission diagram

### 2.4. Post-lockdown scenarios

#### 2.4.1. The worst case scenario and the 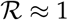 scenario

Most studies concerning the early growth-rate of the epidemic in Wuhan estimated the value of the basic reproduction number to be around 2.0–3.0 (see e.g. [49,52]), also later studies regarding the spread in other countries [18,19] used similar values. Our estimations given in Section 2.2 shows that in Hungary the highest value of the time-varying reproduction number was 2.2, by the Wallinga–Teunis method. Hence, we choose 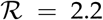 for the baseline reproduction number. Modelling studies [15,16,18,19] highlighted that the worst case, i.e. “do nothing” scenarios lead to an outbreak when the healthcare demand hugely exceeds the capacities at the peak, and the overall mortality can be devastating. Given the current level of preparedness, we do not consider a “do nothing” scenario, and our most pessimistic case assumes that even in the absence of any control measures, a 25% reduction in transmission is realized due to population awareness and behaviour.

On the other hand, the best case is the continuation of the current suppression scenario with 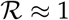, resulting in very small case numbers. However, it is questionable whether it can be sustained until a vaccine is developed and deployed. Below we consider three scenarios illustrating the loss of control for suppressing the outbreak, and assuming a wide community spread of the disease. The efficacy of the mitigation efforts is expressed by a percentage in the reduction of transmission. The primary tool for this is the decrease of contact numbers, but other preventive measures such as hand hygiene or mask wearing may also have an effect in the reduction of transmission.

#### 2.4.2. Weak control: 25% reduction of transmission

Here we consider a weak control of the epidemic assuming there is no centralized control measure introduced, but the number of transmissions is reduced by 25% following a level of behavioural response due to social awareness. Such a reduction decreases the reproduction number to 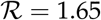. The first column of Figure 6 shows the hospitalization and ICU demand on the top row and the daily incidences on the bottom row as a function of time with the application of this weak control. According to the simulations, in this case, there would be approximately 5.6 million infections with about 22,000 deaths by the end of the outbreak. This suggests that we can expect 57% of the population to gain immunity against the virus and this number is slightly larger than the threshold of herd immunity. At the peak, more than 600,000 people would be infected and there would be a need for more than 7,000 ICU beds and for 21,200 hospital beds at the same time. In this case we would reach the peak in 13 weeks after reopening with such a weak measure. We remark that there is a 20-days window when the daily incidences exceed 100,000, and during this period more than 2.25 million people get infected. In other words, 40% of all the infections occur during these three weeks. For further details see Table 2.

**Figure 6.**
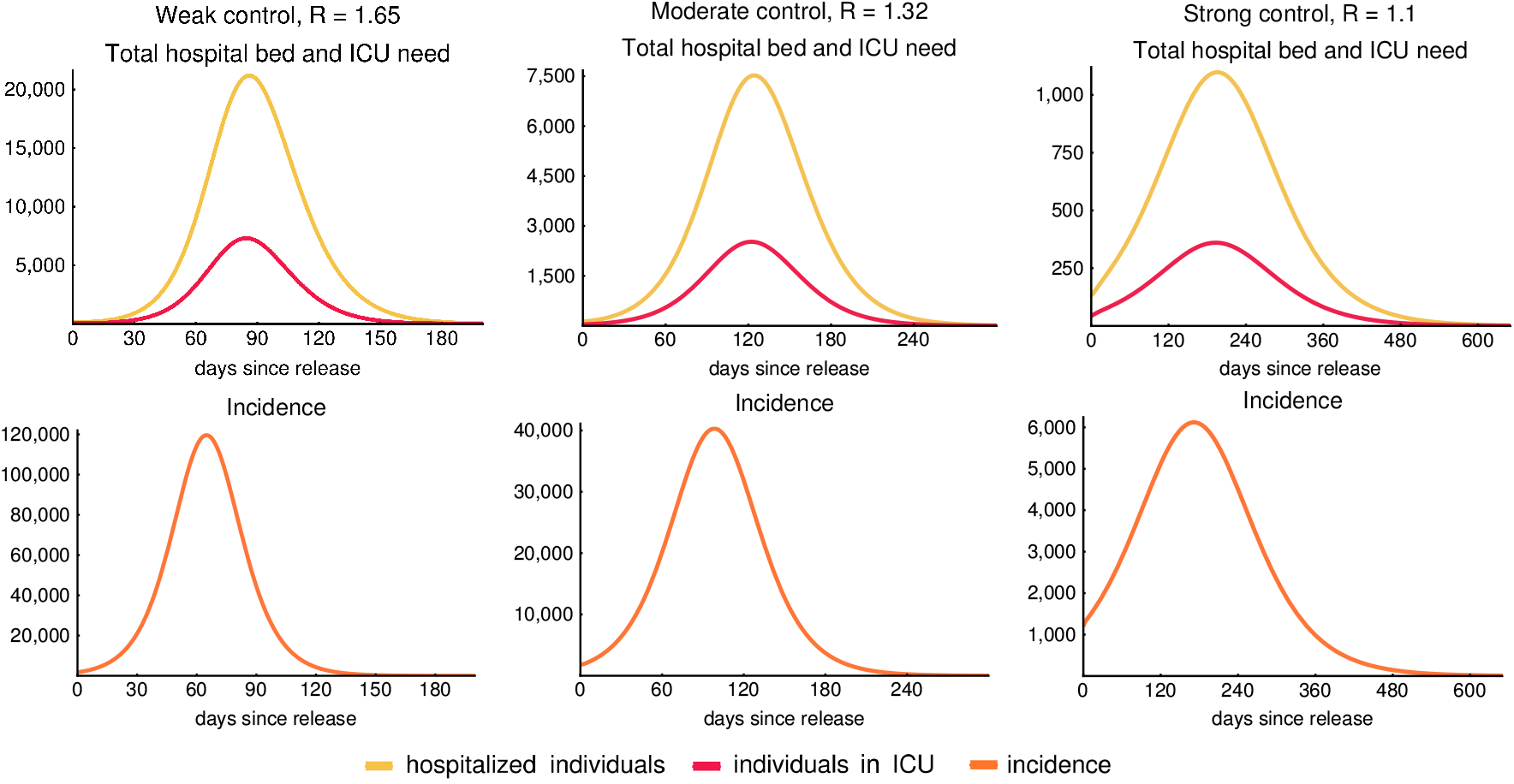
Hospitalization, ICU demand and incidence curves. The figures show the required number of hospital beds (classes *I_h_* + *I_cr_*, yellow) and ICU beds (class *I_c_*, red) need in the first row for 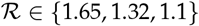 respectively. The second row illustrates the daily incidence (transition from compartment *S* to *L*1 in our model, orange) combining all age groups. Note that the incidence curves peak earlier than the hospitalization curves. The legend at the bottom applies for all figures. Note that the scalings of the figures are different.

**Table 2.** Indicative values of the epidemics in case of the applied control measures. Hospital and ICU bed need at the peak, mortality and the number of recovered people with the expected time it takes to reach 1000 ICU beds is shown in case our control scenarios. See the corresponding time series on Figure 6.

| Transmission reduction | Reproduction number | Hospital bed need at peak | ICU need at peak | Time to reach 1000 ICU beds | Mortality (pers.) | Recovered (of tot. pop.) |
| --- | --- | --- | --- | --- | --- | --- |
| 25% | 1.65 | 21,200 | 7,330 | 7 weeks | 22,200 | 57% |
| 40% | 1.32 | 7,600 | 2,500 | 12 weeks | 12,700 | 36.5% |
| 50% | 1.1 | 1,100 | 350 | - | 4,500 | 14% |
| 60% | 0.9 | - | - | - | - | - |

#### 2.4.3. Moderate control: 40% reduction of transmission

We perform similar simulations for the case of a moderate control, assuming that the reproduction number is decreased to 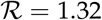 as a result of the control measures. The simulations (third column of Figure 6) show that the number of hospital beds and ICU beds needed is significantly reduced to 7,600 and 2,500 at the peak, respectively. Meanwhile, the daily incidence at the peak is around 40,000. We expect almost 36% percent of the population to be infected throughout the epidemic and gain immunity upon recovery. This is less than required to reach herd immunity. For further information we refer to Table 2.

#### 2.4.4. Strong control: 50% reduction of transmission

In this section, we consider a stronger control achieving a 50% reduction of transmission. This results a decrease of the reproduction number to 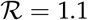. The outcome of this strong control is shown in the third column of Figure 6. A control of such strength significantly reduces the number of all infected and hospitalized cases and of those needing intensive care treatment. The number of required intensive care beds (around 350) is far below the available capacity even at the peak of the epidemic and also the number of hospital beds needed is reduced to a rather low level – around 1100 at the peak. The total number of fatalities in this scenario is about 4500. Meanwhile the epidemic would last for more than a year and the cumulative number of all infected remain far below the level of herd immunity threshold, so we can expect further outbreaks when the measures are relaxed.

### 2.5. Age-specific issues

Several key parameters of the model are highly dependent on age. Intervention strategies and the relaxation of various measures have to take into account the fact that different age groups have different risks and different roles in the transmission.

Although the number of children infected with COVID-19 has been reported worldwide relatively small in comparison with other age groups [53], some evidence shows that children and adolescents may become infected and spread the disease as other age groups [54]. Moreover, children and adolescents usually have a high number of contacts. Thus, school closures can be expected to be an efficient tool to reduce the contacts and transmissions. Besides school closures, it is important for younger individuals to avoid meeting older and other high risk people.

Elderly people have a higher chance of developing symptoms, and a higher percentage of them needs hospitalization and intensive care, hence these groups need more protection. Age-specific interventions include avoiding contacts with elderly by providing special time slots for shopping, in post offices etc., or closing/reopening schools.

Introduction of various age groups in our model enables us to study such age-specific interventions and analyse their direct and indirect effects on all groups. On the stacked diagrams of Figure 7 we present the contributions of the age groups to the mortality and the number of recovered individuals. Columns of this figure show the effect of the weak, moderate and strong control that we previously discussed in details in Section 2.4 and Table 2. Here we would like to emphasise that in case of each control measure the most vulnerable age groups are the groups of elderly (60–69, 70–79, 80+) people as they suffer most of the fatalities, meanwhile they are predicted to produce only a small fraction of the cases in the population.

**Figure 7.**
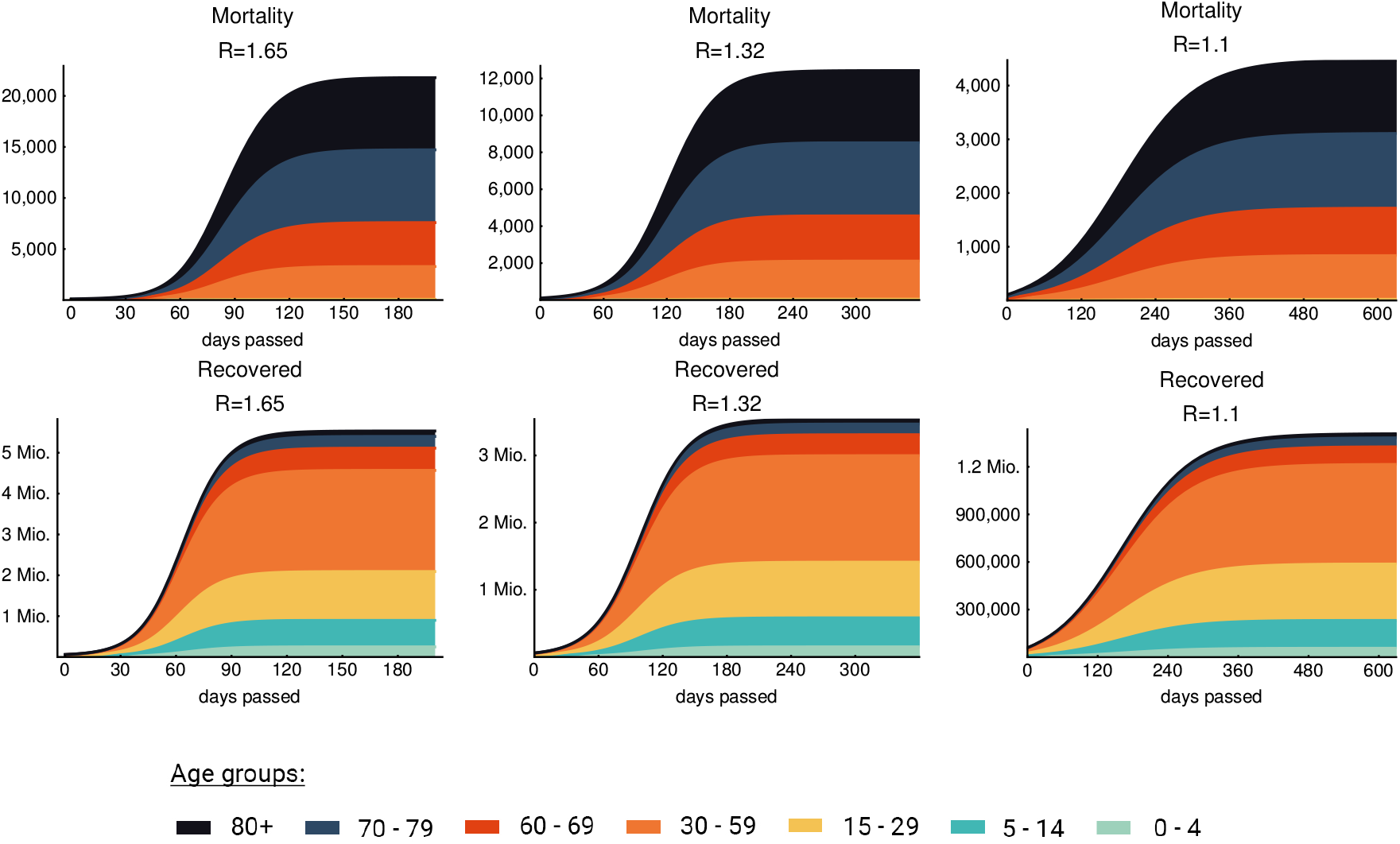
Age-specific mortality and recovery. The figure shows the effect of the weak, moderate and strong control (25%, 40% and 50% general contact reduction respectively). Every age group covers at most one decade except the group of “middle aged” that contains three decades. According to our model elderly people (60+) are predicted to produce most of the fatality cases in each scenario. The legend on the bottom applies for all figures.

#### 2.5.1. School closures

School closure is taken into account by omitting the school component of the contact matrix and halve the *other* contacts [55] of children and young adults (between age 0–29), which provides a new global contact matrix for this intervention. We also incorporate the weak control (25% general decrease in transmission, cf. Section 2.4.2) to this scenario.

Figure 8 shows that this measure decreases the hospital bed and ICU needs to approximately 50% compared to the case when we only apply weak control. Moreover, closing schools postpones the peak of the epidemic (by about one month in case of the above setting), suggesting that children may play a significant role in transmission due to their large number of contacts, even though they give negligible contribution to the overall mortality, cf. top row of Figure 7).

**Figure 8.**
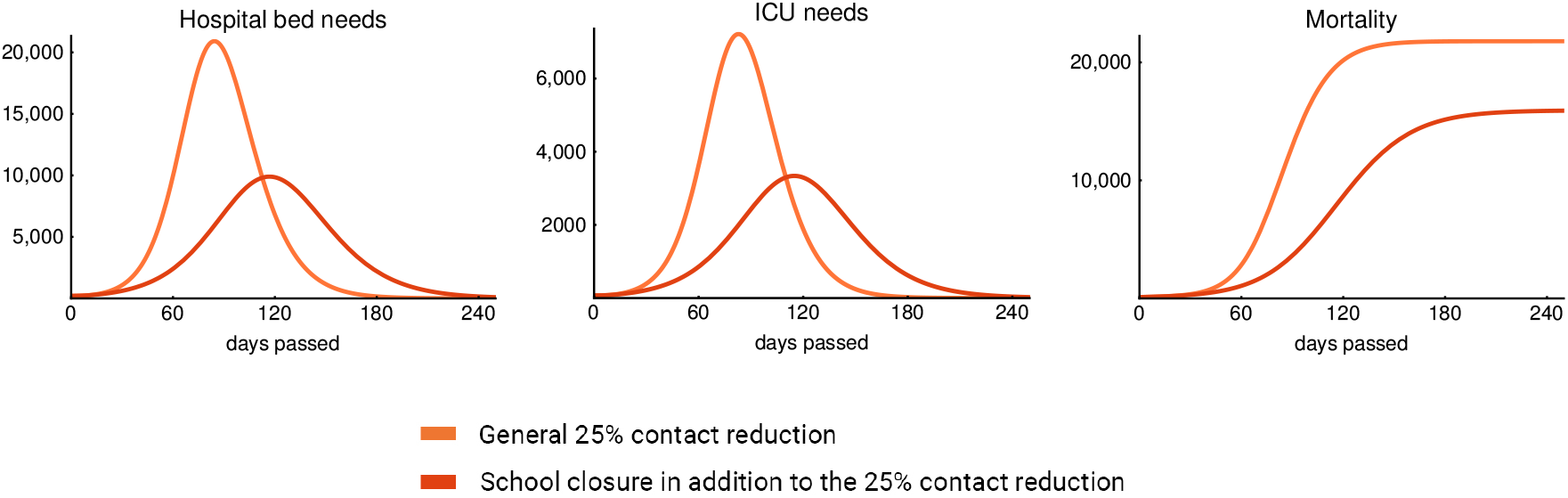
Effect of school closure. Simulations suggest that school closures – if maintained for a long period – effectively decrease hospital bed and ICU needs and significantly postpone the peak of the epidemic.

The effect of school closure combined with the 25% general reduction in transmission is comparable with the effect of the moderate control (40% reduction in transmission, cf. Section 2.4.3) regarding the hospital bed and ICU need, but not as significant as the moderate control in decreasing the mortality (Figure 7 middle column). However, to achieve this, schools need to be closed for an extended period of time, which may not be feasible. We also point out that a stand alone closure of preschools and primary schools is not sustainable without a certain amount of home office of the parents, but this opens up sociological and economical questions that we do not address here.

#### 2.5.2. Elderly protection

The elderly being the most vulnerable group of the population, when it comes to relaxation of measures introduced against the spread of COVID-19. Most countries handle these age groups separately from the rest of the population, e.g. separate time slots for shopping continue to exist and elderly are encouraged to keep the same level of social distancing. To include these effects in our model, we manipulate the entries of the contact matrix involving older age groups separately from the remaining parts.

Figure 9 illustrates that in addition to the weak control, if 50% and 100% reduction of the outside household connections of elderly people is applied, then we can expect about 25% and 50% reduction in the hospital, ICU bed needs and mortality. The epidemic curves only slightly shift to the right suggesting that elderly people do not play important role in the transmission of the disease due to their low number of contacts. 100% reduction of contacts outside the household is again not feasible, as this would mean the complete isolation a large sub-population. We plotted this scenario only to show the theoretical limits of this approach.

**Figure 9.**
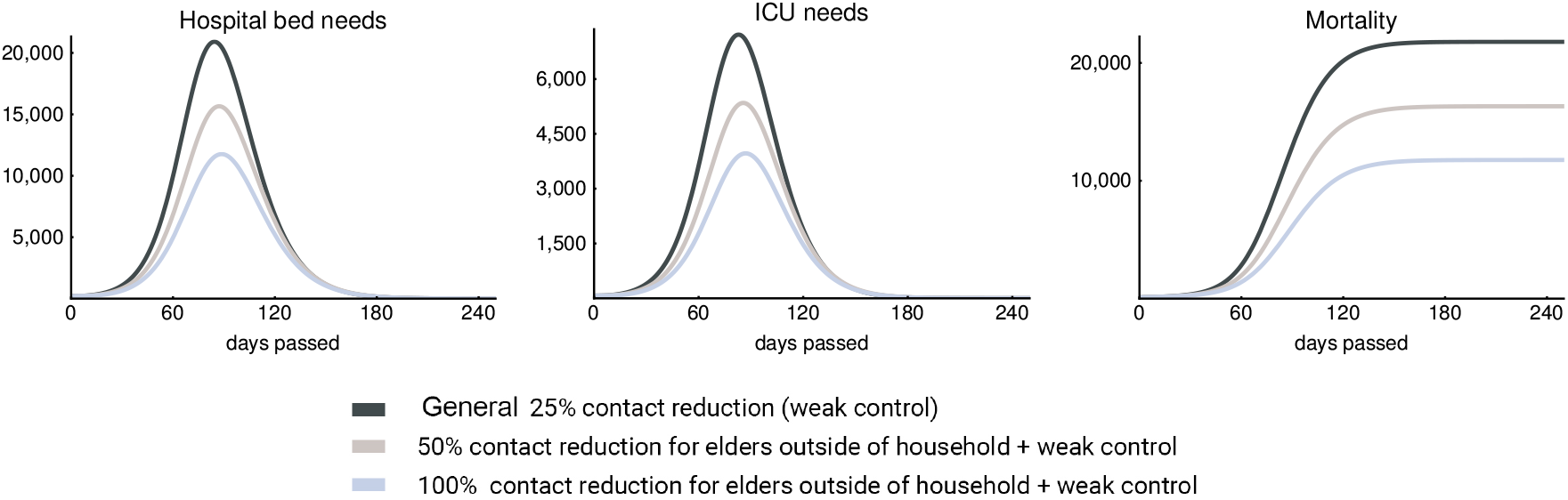
Elderly protection. Figures show the effect of an additional contact reduction of elderly people in case of a weak control. The figures suggest that the selective protection of elderly people can successfully reduce the peak ICU need and the overall mortality, yet it has a theoretical limit.

### 2.6. Role of seasonality

In this section, we consider the impact of seasonal effects. Seasonality of respiratory viruses can be attributed to a combination of factors, including the survival of the virus in different environmental conditions, changes in contact patterns (such as school holidays), less time spent in closed spaces where the highest number of transmissive contacts are made, and potentially seasonal changes in the health conditions of the population as well. To express this behaviour, we define a time-dependent parameter

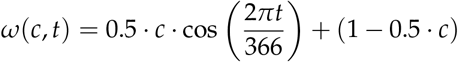

by which we scale the transmission rate *β*. Parameter *c* denotes the magnitude of the effect of seasonality on the number of contacts. We investigate the epidemic curves in case of the weak, moderate and strong control with seasonality parameter *c* ∈{0, 0.1, 0.2, 0.3}. During the summer, these values of *c* eventuate a 10%, 20%, and 30% further decrease in transmission as that is when the seasonality curve attains its minimum. The case *c* = 0 means there are no seasonal effects at all, while *c* = 0.3 is a strong seasonality similar to H1N1 [56]. See the top left image of Figure 10 for the seasonality functions *ω*(*c*, *t*) corresponding to the different *c* values.

**Figure 10.**
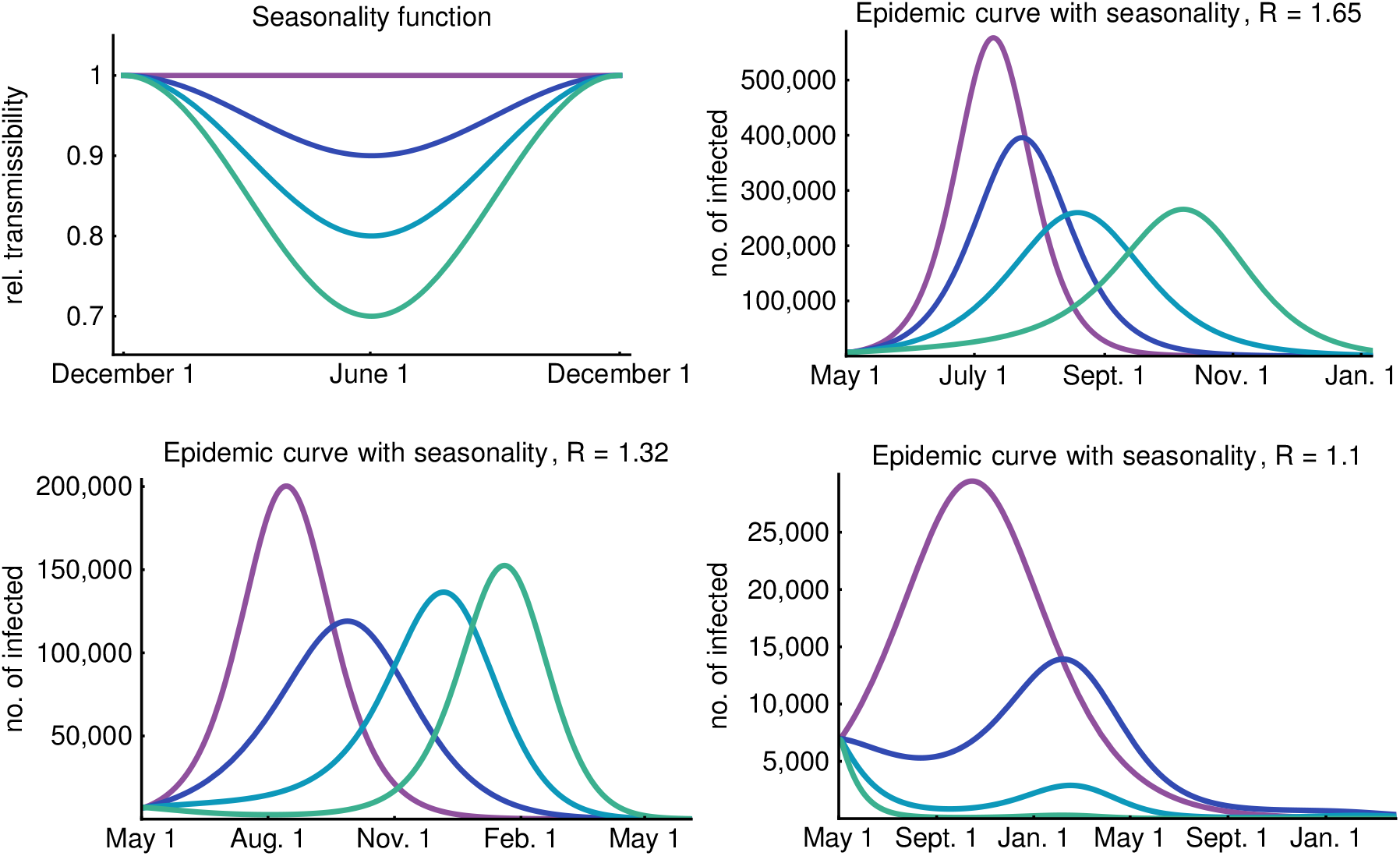
Top left figure shows the relatively transmissibility of the virus during the year. In the other figures, the number of infected individuals is shown with 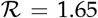,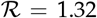 and 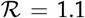. Purple denotes no seasonality (*c* = 0), blue curves correspond to weak seasonality (*c* = 0.1), turquoise curves correspond to moderate seasonality (*c* = 0.2) and green curves correspond to strong seasonality (*c* = 0.3).

As we have seen in Section 2.4, decreasing the reproduction number decreases and postpones the peak of the epidemic curves. Seasonality causes a similar delay in the peak of the epidemic due to decreased transmission rates in the summer months. Counter-intuitively, it cannot be said in general that stronger seasonality leads to a smaller peak (cf. bottom left image of Figure 10). The reason for this is that the impact of seasonality is not only determined by the decrease in the transmission rate but the temporal relation between the peak of the epidemic and the minimum of the seasonality function is also an important factor. This phenomenon is well illustrated in Figure 10 where three scenarios (weak, moderate and strong control) are presented along with the assumed seasonality functions for the aforementioned values of *c*.

In the upper right image of Figure 10, corresponding to a weak control, one can observe that increasing the effect of seasonality first decreases the peak, but after a certain value (*c* = 0.3 in our example) the epidemic is so much suppressed in the summer months that the peak shifts to the right and even slightly increases in winter months compared to the *c* = 0.2 scenario.

For the case of moderate control, shown in the lower left figure, this effect is much more significant. Note that the peak of the epidemic (without seasonality) is so far from summer (the minimum of the seasonality curves) that increasing the effect of seasonality results in a significantly higher peak. It can be seen that strong seasonality eventuates a long “plateau” phase when the epidemic curve does not increase in a period of 6 months. During this time, only a small fraction of the population goes through the infection and a massive number of susceptibles remain in the system, only to get infected a few months later. This phenomenon is responsible for the increased peak of *c* = 0.3 compared to the *c* = 0.2 case.

The lower right figure shows that the reduction of transmission during the warm months together with a strong control can decrease the number of infected in such an extent that the peak, even if arriving in the winter months, is significantly smaller.

A general observation is that seasonality is the most beneficial if the peak of the epidemic is close to the summer months. Of course, this is highly dependent on the starting time of the outbreak.

### 2.7. Spatial heterogeneity

Hungary is a relatively small country, however, significant differences were observed between regions in the reported case numbers. The capital, Budapest has 1,75 million inhabitants and further 1,23 million people live in its surrounding Pest county. Budapest and Pest county are highly connected by commuters with connections to other regions as well. The high connectivity of the capital with other countries contributed to the earlier appearance of the disease in Budapest, and most of the cases were reported from this central region of the country.

To address the role of spatial heterogeneity in the evolution of the epidemic curve, in addition, we considered a metapopulation model as well, where the population is distributed among patches, representing geographic regions of the country. For the sake of simplicity, here we only present results from a two-patch model, separating Budapest and Pest county (patch 1, population of approx. 3,000,000) from the remaining parts of the country (patch 2, population 6,800,000).

We assumed different transmission parameter *β* for each patch. Based on Hungarian mobility data, we assumed 400,000 daily travels between the two patches in case of normal circumstances and investigated the effect of the lockdown of Budapest and the surrounding Pest county by decreasing the number of daily travels to 10,000. We considered the contact matrix for both patches to be the same as in the homogeneous model described in Appendix C. The biological and medical parameters are assumed to be the same in each patch but the local reproduction number may differ, as well as the age structure of the population. For obvious reasons, individuals in compartments 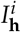, 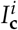, 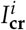 and *D^i^* do not travel. Let *travel***_p,q_** denote the number of travels from patch *p* to patch *q*. To derive travel rates *t***_patch,q_** for each age group *i*, we divide the number of travels with the population of the appropriate patch:

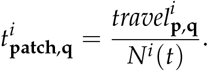

The left-hand side of Figure 11 illustrates that the two-patch model reproduces the uniform model in case we use the same 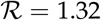 for both patches as well as for the uniform model and we assume 400,000 daily travels between the patches. The middle figure shows that the uniform model slightly overestimates the size of the epidemic as the peak of the aggregated two-patch model is smaller than that of the uniform model in case 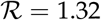 remains the same and we reduce the daily travels to 10,000 corresponding to the separation of Budapest and Pest county from other regions. Although the epidemic curves of the patches are shifted, the aggregated result shows that this setup does not provide significantly different dynamics. Lastly, on the right-hand side of Figure 11, we further investigate the scenario of 10,000 daily travels, and choose the local reproduction numbers of the patches to vary around 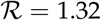, namely, we take 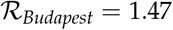 and 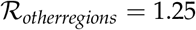. These values were selected to reflect the higher population density of the capital, proportionally to the population in the two patches. Due to the difference in the local reproduction numbers, we may observe an increased number of cases in Budapest with an earlier peak and fewer infections in other regions.

**Figure 11.**
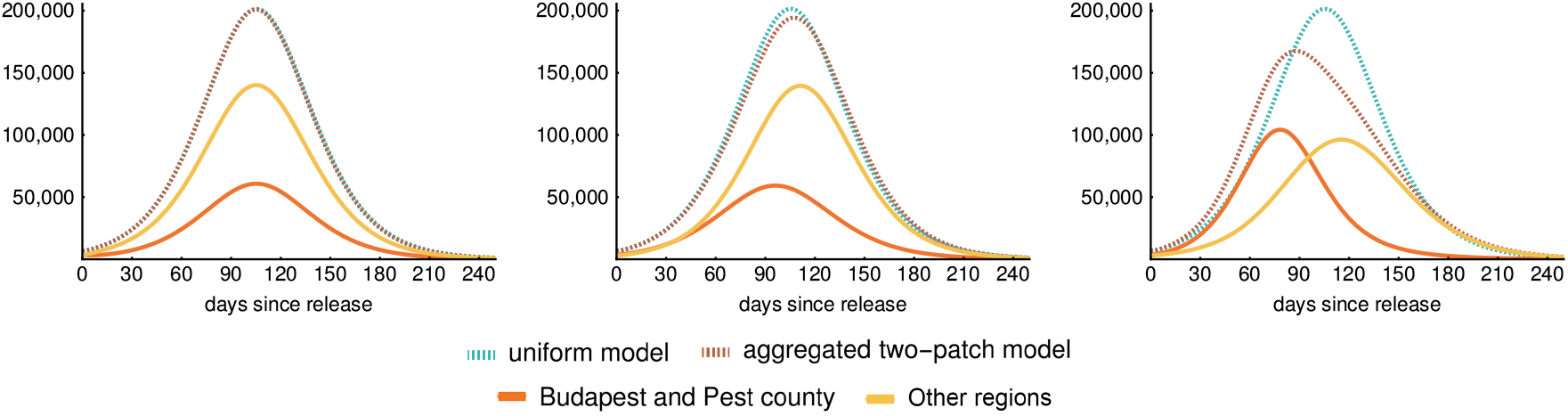
Epidemic curves of the regions: sum of the infective compartments. (*I_p_*, *I_a_*,_1_, *I_a_*_,2_, *I_a_*_,3_, *I_s_*_,1_, *I_s_*_,2_, *I_s_*_,3_). The first figure shows that the two-patch model reproduces the uniform model for equal 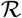 and no travel reduction. The middle figure shows the uniform model slightly overestimating the size of the epidemic for reduced travel rates and equal 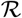. The last figure shows an increased number of cases and earlier peak in Budapest, while less infections in other regions, for different 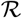 in the two patches and large travel reduction.

### 2.8. The impact of implemented measures since mid-March

The most important implemented measures are summarized in Table 3. To assess their impact, we compared the reported case numbers adjusted by the ascertainment rate 1:17 to the simulated outbreak curve with 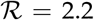 (Figure 12 on the left, logarithmic scale). Here we assumed that the ascertainment rate did not change in time, which may not be the case. One can see that the epidemic was on the 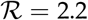 trajectory, which could have been devastating if continues. The data shows a clear deviation from this scenario early April, two weeks after strict social distancing started. The slope of the epidemic curve further decreased mid-April, following the stay at home measures by two weeks.

**Table 3.** Measures applied in Hungary with date of introduction.

| Date | Measure | Reported number of cases at time of introduction |
| --- | --- | --- |
| March 8 | Banned visits to health care institutions and long-term care facilities | 9 |
| March 9 | Suspension of Northern Italy flights | 12 |
| March 11 | Emergency notification | 16 |
| March 12 | University closures, no entry for non-Hungarian passengers to Hungary from Italy, China, Korea and Iran | 19 |
| March 16 | School closures | 50 |
| March 17 | Shortened opening time of shops, ban on events | 58 |
| March 28 | Stay at home measures | 408 |
| May 4 | Partial lifting of stay at home measures and opening of restaurants in the countryside (except Pest county where from May 14) | 3065 |
| May 18 | Lifting of stay at home measures and opening of shops and outdoor areas of restaurants in Budapest | 3556 |

**Figure 12.**
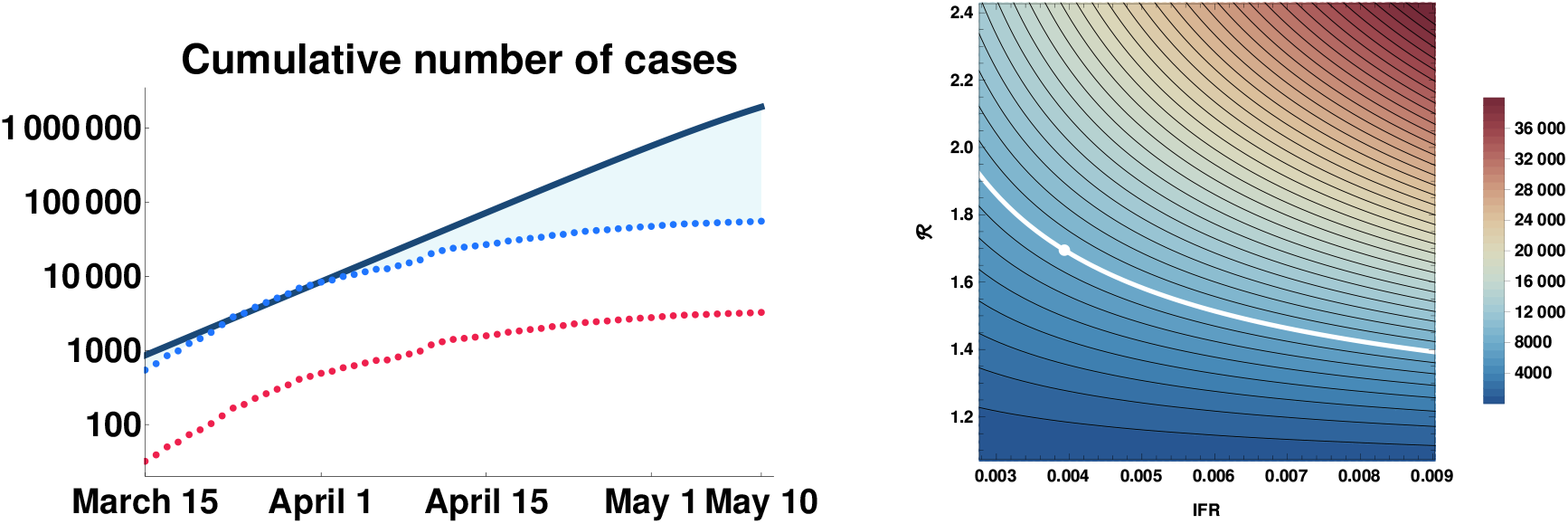
Left: the impact of control measures on the epidemic trajectory. Red dots are cumulative numbers of reported cases, blue dots are corrected data by underascertainment rate, solid curve is simulated cumulative numbers with 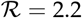 and the absence of measures. Right: Sensitivity of the peak ICU demand to transmissibility and severity of COVID-19. Top right corner is similar to the worst case scenario of [18]. The white dot is our most pessimistic scenario (weak control).

Overall, due to the compliance of Hungarian society with the social distancing measures, around half million infections were averted by the end of April, compared to the "do nothing" scenario, which could have reached 1-2 millions in May if further doublings would have been allowed.

### 2.9. Parameter uncertainty, sensitivity and other limitations

Our work has several limitations. Due to limited testing and the large number of asymptomatic and mild cases, there was a huge uncertainty in the number of true cases, especially in the early weeks. Now, with the help of [46] we will have a good estimate of the overall ascertainment rate over this period, but it is still unclear how this rate evolved in time. The transmission model has the weaknesses that all compartmental models have: we assume homogeneous population apart from the age structure. We added some further heterogeneity in space (patch model) and time (seasonality). In our scenarios, we assumed a constant reduction in transmission, while in reality the control measures and the behaviour of the people were continuously changing. Hence, such scenarios can not be considered as predictions, as we can not expect such unchanging circumstances for months. The role of children in this pandemic is still not clear, in our modelling we assumed that they are equally susceptible, and equally infectious once they develop symptoms, but we used an age-specific probability for developing symptoms.

The model has a large number of parameters, many of those have uncertainty. The most important ones in regard to the burden on the healthcare system are hospitalization rates, probability of intensive care need, mortality, all of those depending on age. We do not have too much data for this from Hungary, hence we used parameters taken from the literature. A full sensitivity analysis is beyond the scope of this study, but we present a sensitivity chart for a crucial output of an outbreak, which is of concern in many countries: the peak ICU demand, including the need for mechanical ventilators, to assure that all patients receive the necessary care, and mortality will not skyrocket because of the overwhelmed healthcare system. This was one of the key questions in other modelling studies.

For an uncontrolled epidemic in the UK, [18] estimated a peak in ICU bed demand more than 30 times greater than the maximum capacity in these countries. In a study for the United States, [19] projected that at the outbreak peak, 3 times more ICU beds would be needed than the total number of ICU beds in the US, and 85% isolation of cases reduces the demand for ICU beds to the normal capacity. In the Île-de-France region, [16] estimated that the peak number of ICU beds needed would exceed more than 40 times the regional capacity if no strategy is implemented after lockdown, and only efficient case-finding and isolation applied parallel with social distancing could decrease ICU demand below the maximum capacity throughout the epidemic. For Australia, [15] studied three capacity expansion scenarios (2, 3 and 5 times expansion, respectively), and even in mitigated scenarios, demand is estimated to be higher than the number of available beds. Additional social distancing measures were shown to reduce the epidemic to a level where a reasonable expansion of ICU capacity can be sufficient.

The peak ICU demand crucially depends on two factors: the probabilities of developing sever disease, and the shape (in particular the peak size) of the epidemic curve. We plotted a heatmap of the peak ICU demand in Figure 12, compiled from hundreds of numerical simulations. Transmissibility (vertical axis) is expressed by the reproduction number 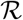. Disease severity, for simplicity, is expressed by the IFR. In fact, here we used a scaling factor for the probability of hospitalization, with the baseline corresponding to the parameters in Table B.3. In our weak control scenario (Section 2.4.2), the infection fatality rate is 0.4%, which is a bit lower than the finding of [46]. However, during the first wave in Hungary the schools were closed and COVID-19 disproportionately affected the vulnerable population. In our scenarios we assume a widespread community spreading, hence younger generations appear in higher numbers, thus the infection fatality rate is expected to be smaller. In any case, by the scaling of the hospitalization rate (while leaving the probability of intensive care and fatal outcome given hospitalization intact), we explored a wider range of IFRs. We found that indeed the peak ICU demand can vary across a large interval. From the shape of the level curves in the heatmap, we can conclude that the peak ICU demand is more sensitive to 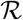 than to the IFR, hence flattening the curve is indeed of utmost importance to avoid exceeding healthcare capacities.

## 3. Results

The first COVID-19 case was detected, laboratory confirmed and then reported through the Hungarian Notifiable Disease Surveillance System on March 4, 2020. Well tailored, effective, combined non-pharmaceutical control measures have been introduced promptly in Hungary in the very early phase of the outbreak (see Table 3), accompanied with a high level of compliance for social distancing. Online surveys [51] and indirect data (such as traffic data, passenger volumes on public transportation etc.) all showed a drastic reduction in the number of contacts and mobility. Accordingly, the Hungarian epidemic curve was strongly suppressed. As of May 10, 2020, the cumulative number of reported confirmed COVID-19 cases were 3284 (33.1 cases per 100,000 population), including 421 deaths. The epidemic peaked on April 9 with 209 newly reported cases. SARS-CoV-2 was not able to sustain long transmission chains in the community, however, it was able to cause outbreaks mostly in healthcare institutions and long-term care facilities: nearly two third of the reported cases are connected to such institutions. The proportion of cases in health care workers gradually increased during the epidemic. They had tenfold risk to become confirmed COVID-19 cases compared to the general population. Due to effective measures, the virus could not spread significantly from closed communities and health care workers to the wider population. The age specific case fatality rate showed a similar pattern to other countries: of the 421 deaths reported by May 10, 375 (89.1%) belonged to the 65+ age group.

We tracked the temporal variation of the effective reproduction number real time, which showed a steadily decreasing trend, interrupted by an outlying outbreak in a long term care facility. We identified the time intervals when the effective reproduction number was below or around the critical threshold 1. The adjusted case fatality ratio was also estimated real-time, and well predicted the eventual case fatality ratio in one month advance. Benchmarking the CFR to other countries, we estimated underdetection rate to be 10-20 times, and the true cumulative number of COVID-19 cases to be between 32,840 and 65,680. These results are consistent with data from the preliminary results of a large scale seroepidemiological survey, carried out in Hungary in May 2020, where the seroprevalence of SARS-CoV-2 infection was estimated to be between 22,399 and 92,624 [46]. Based on these data and the number of reported cases, underdetection is likely to be between 6.8–28.2, and true CFR may be lower than 1.5%, and the IFR is roughly half of that.

As control measures are being successively relaxed since May 4, we established an age-structured compartmental model to investigate several post-lockdown scenarios, and projected the epidemic curves and the demand for critical care beds assuming various levels of sustained reduction in transmission. Special measures designed to reduce the contact number of the elderly population as well as school closures can reduce the peak hospital bed demand and the overall mortality, however these measures also have their limitations. A metapopulation version of the transmission dynamics model has also been studied, and we reported some results for a two-patch case, where the Budapest region is considered separately from the rest of the country. Due to the high connectedness, the epidemic curves of the two-patch system are not much different from the spatially homogeneous case. To achieve a noticeable reduction in the overall peak size due to spatial heterogeneity (where the local peak times are shifted in the regions), a large reduction in the mobility rates is necessary.

Since the vast majority of the population is still susceptible, a weak or even a moderate reduction in the transmission, compared to the baseline, could result in a large second outbreak with significant mortality and high peak ICU demand. Therefore, high level of alertness needs to be maintained to avoid such scenarios.

The seasonal behaviour of SARS-CoV-2 is not completely understood yet, thus we considered a range of possibilities from the absence of seasonality to strong seasonality similar to H1N1. The interplay of seasonal effects with the post-lockdown contact numbers can generate a variety of disease dynamics, thus a confident forecast of the timing and the size of a potential second wave is not possible at the moment.

The effectiveness of strict social distancing measures, such as school closures and stay at home measures with good compliance is likely to be very high, however such interventions have devastating consequences on the society and on the economy, thus not sustainable on long term. Modelling results [57,58] suggest that combined multiple interventions, including moderate contact decrease, high COVID-19 detection rate, effective contact tracing and good compliance with personal protective instructions, may have substantial impact on transmission, and are able to keep the reproduction number around one.

## Data Availability

The developed software package is available from the link displayed in the manuscript.

https://github.com/zsvizi/covid19hun

## Acknowledgement

This work was done in the framework of the Hungarian National Development, Research, and Innovation (NKFIH) Fund 2020-2.1.1-ED-2020-00003. Some authors were also supported by EFOP-3.6.1-16-2016-00008 (Zs.V.), NKFIH KKP 129877 (F.B.), NKFIH FK 124016 (T.T.), NKFIH PD 128363 (A.D.), Bolyai Scholarship of HAS (A.D.), TUDFO/47138-1/2019-ITM (F.B., P.B.).

## Technical Appendices

The Appendices contain the governing equations of our transmission model in Appendix A. We detail the parameters in Appendix B that we have used to tune the system. Finally, Appendices C and D present the utilized contact matrix and the computation of transmission rates via the next generation matrix, respectively. The codes were implemented in Wolfram Mathematica and available at https://github.com/zsvizi/covid19hun.

## Appendix A The governing equations of the transmission model

The governing equations of the transmission model described in Section 2.3 take the form

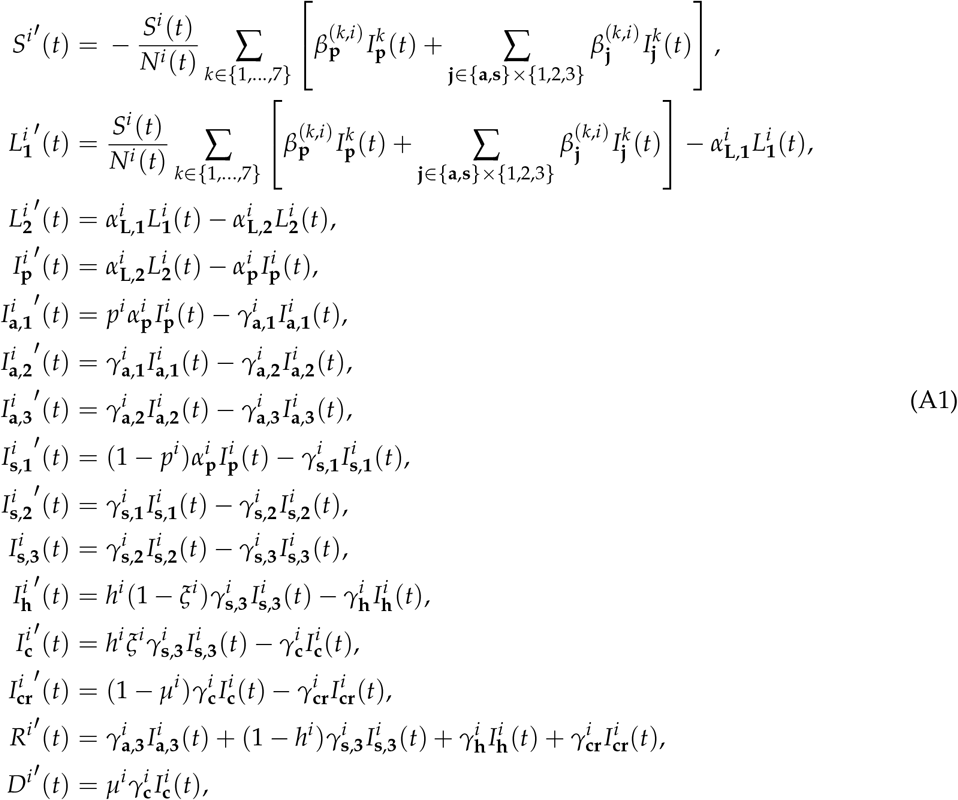

where the index *i* ∈{1,. . .,7} represents the corresponding age group.

Governing equations of the metapopulation model where *p*_1_ ∈{1,2,. . .,#patches} are

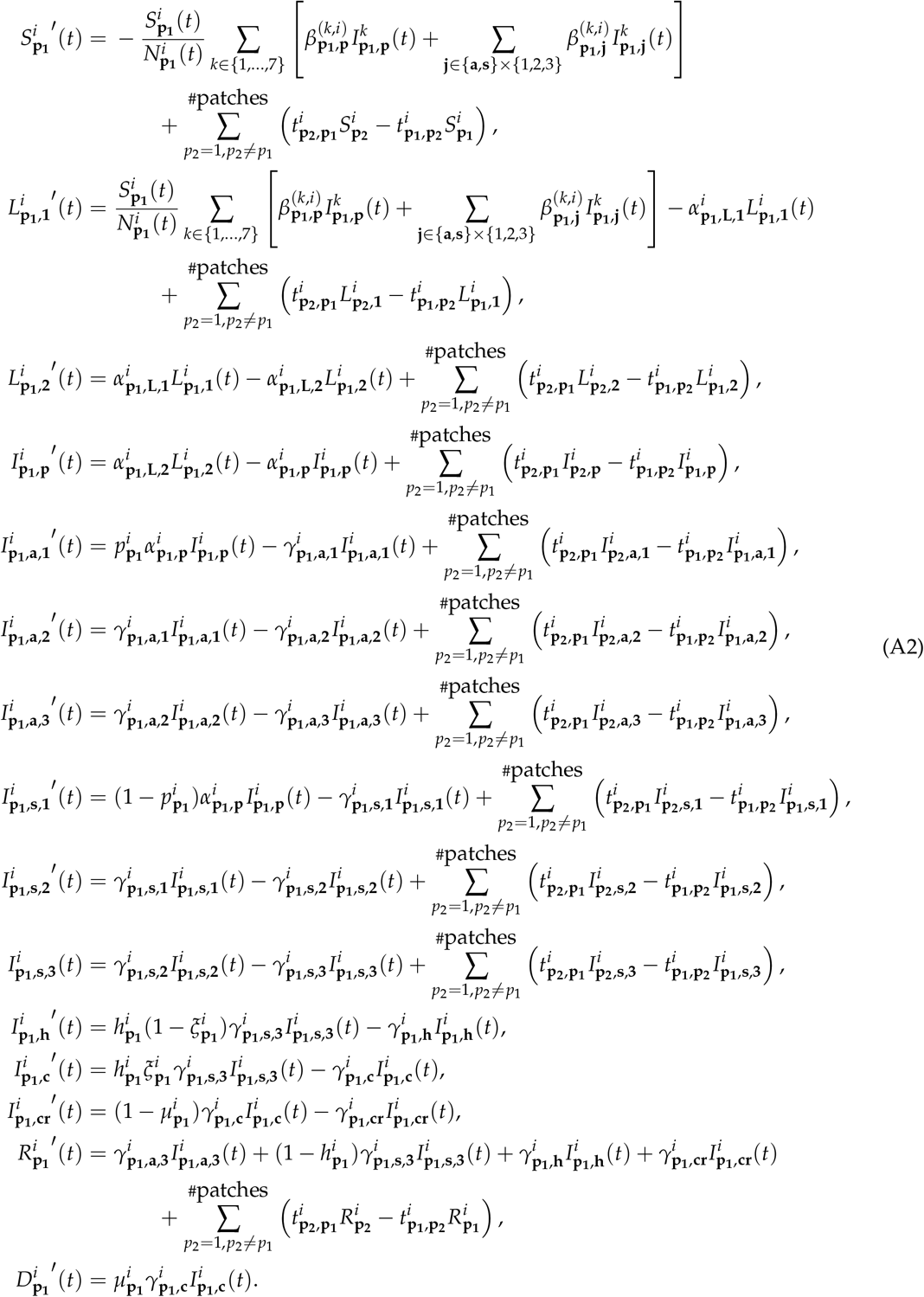

## Appendix B Model parameters

We have chosen our model parameters based on comprehensive literature review and present them here, except the transmission rates 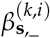 which are left for Part D. For the incubation period we assume hypoexponential (generalized Erlang) distribution with parameters (1.6, 1.6, 2). This way, the average incubation period is 5.2 days: the same length and very similar shape of the probability distribution function was estimated in [49], and this distribution has the observed concavity properties as well (see [59]). Also, this estimate is consistent with [40], and such values have been used in [15,16,18,19]. The first 3.2 days are the latent period [15] and the past 2 days are the presymptomatic period [15], when transmission is already possible with similar rate as at symptom onset [48]. Therefore, we use the same transmission rates for the presymptomatic and symptomatic infectious periods. For the transmission rate of asymptomatic infected individuals, we use a reduction factor 0.5 [16,19,47].

For the length of infectious periods (both symptomatic and asymptomatic), we assume a gamma distribution with Erlang parameter 3 (coherent with the SARS study [50]), and an average length 3 days of infectivity. Although full recovery and viral shedding may take much longer, the infectiousness throughout the course of infection is mostly concentrated to this period [48,60]. The choice of 3 days is also justified by [48] and [61], who estimated that around 40% of transmissions occur during the presymptomatic period, and it is also within the range of infectious periods used by [16] and [19].

The average stay in hospital is assumed to be 10 days, in accordance with the seven days median reported in [62] using over 16000 patient’s data in the UK. Similarly, the average duration of critical care is assumed to be 10 days, in accordance with the ICNARC report [63]. Very similar numbers were reported in the US [64], and were used in other modelling studies [15,18,19]. For those who recover from intensive care, we assumed a 14 days hospitalized rehabilitation period.

The periods above associated to the average time an individual spends in each compartment over the course of the infection are age-independent and summarized in Table B1.

**Table B1.** Age-independent epidemiological parameters of COVID-19 Assumed to be valid for all age groups *i* ∈{1,. . .,7}. References and explanations are in Appendix B.

| Duration of |  | Value |
| --- | --- | --- |
| Incubation period | $(\alpha_{L,1}^i)^{-1} + (\alpha_{L,2}^i)^{-1} + (\alpha_p^i)^{-1}$ | 5.2 days |
| Latent period | $(\alpha_{L,1}^i)^{-1} + (\alpha_{L,2}^i)^{-1}$ | 3.2 days |
| Presymptomatic (infectious) period | $(\alpha_p^i)^{-1}$ | 2.0 days |
| Infectious period of $I_a^i$ | $(\gamma_{a,1}^i)^{-1} + (\gamma_{a,2}^i)^{-1} + (\gamma_{a,3}^i)^{-1}$ | 3.0 days |
| Infectious period of $I_s^i$ | $(\gamma_{p,1}^i)^{-1} + (\gamma_{p,2}^i)^{-1} + (\gamma_{p,3}^i)^{-1}$ | 3.0 days |
| Hospitalization | $(\gamma_h^i)^{-1}$ | 10.0 days |
| Intensive care |  |  |
| until transition to R or $I_{cr}^i$ | $(\gamma_c^i)^{-1}$ | 10.0 days |
| Recovery in $I_{cr}^i$ | $(\gamma_{cr}^i)^{-1}$ | 14.0 days |
| Relative infectiousness |  |  |
| Presymptomatic vs Symptomatic | $\beta_p^{(k,i)} / \beta_{s,-}^{(k,i)}$ | 1.0 |
| Asymptomatic vs Symptomatic | $\beta_{a,-}^{(k,i)} / \beta_{s,-}^{(k,i)}$ | 0.5 |

Next, we discuss the age-specific parameters, which are mostly related to the outcome of infections. We stratified the population into the following seven age groups: 0–4, 5–14, 15–29, 30–59, 60–69, 70–79, 80+ years old. Using the data from the Hungarian Central Statistical Office (KSH), we obtain the division shown in Table B2.

According to [65], a fraction 0.8 of infected children (under 18 years old) are asymptomatic or mild cases. This value was used in [19] as well. We set the probabilities of the infection following mild or asymptomatic course in an individual according to Weitz *et al*. [47].

**Table B2.**
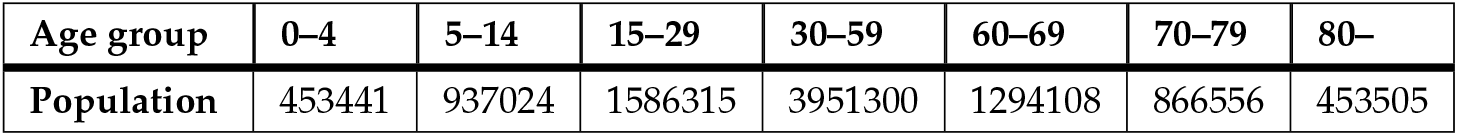
Age groups of the Hungarian population

| Age group | 0–4 | 5–14 | 15–29 | 30–59 | 60–69 | 70–79 | 80– |
| --- | --- | --- | --- | --- | --- | --- | --- |
| Population | 453441 | 937024 | 1586315 | 3951300 | 1294108 | 866556 | 453505 |

The probabilities of hospitalization given infection *h^i^* and of requiring intensive care in addition *ξ^i^* are based on the work of Moss *et al*. [15]. The ratios of fatal outcomes *µ^i^* are derived from the ICNARC report [63] comprising 6720 ICU case reports from UK. All these age-dependent parameters are listed in Table B3.

**Table B3.** Age-dependent epidemiological parameters of COVID-19

| Probability / Age group |  | 0–4 | 5–14 | 15–29 | 30–59 | 60–69 | 70–79 | 80– |
| --- | --- | --- | --- | --- | --- | --- | --- | --- |
| Asymptomatic course | $p^i$ | 0.95 | 0.8 | 0.7 | 0.5 | 0.4 | 0.3 | 0.2 |
| Hospitalization or intensive care (from $I_{s,3}^i$) | $h^i$ | 0.00045 | 0.00045 | 0.0042 | 0.0442 | 0.1162 | 0.2682 | 0.4945 |
| Intensive care (given hospitalization) | $\zeta^i$ | 0.333 | 0.333 | 0.297 | 0.294 | 0.292 | 0.293 | 0.293 |
| Fatal outcome (from $I_{cr}^i$) | $\mu^i$ | 0.2 | 0.2 | 0.216 | 0.3 | 0.582 | 0.678 | 0.687 |

## Appendix C Contact matrix

For creating our contact matrix *M*_cont_ we have utilized the work by Prem, Cook and Jit [55]. As we have divided the Hungarian population into seven age groups (sixteen in Prem *et al*.), we have aggregated the corresponding values into the 7 × 7 contact matrix

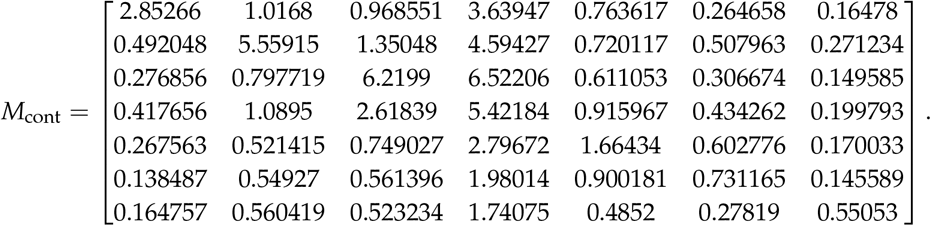

For more insight, we include its heatmap in Figure C1.

## Appendix D Transmission rates and the next generation matrix

Recall that we have assumed presymptomatic patients, that is members of classes 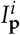, to be as infectious as symptomatic patients. In addition, patients with no or mild symptoms (those in 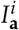) possess a transmission coefficient half of the baseline.

Thus, our task is to give reasonable estimates for the rates 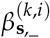 corresponding to the transmission rate of the symptomatic individuals from age group *k* to group *i*. To that end, we follow the terminology and techniques of [66] to compute the Next Generation Matrix (NGM) and the baseline transmission rate *β*_0_. Finally, the desired coefficients are obtained by taking into account the relative contact rates between age groups via the contact matrix presented in Appendix C.

First, let us consider the infectious subsystem of (A1), namely, equations describing 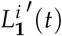, 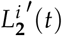, 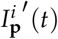, and 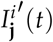 with **j** ∈{**a**, **s**}×{1, 2, 3}, *i* ∈{1,..., 7}. Linearizing this w.r.t. the disease free equilibrium yields the linearized infectious subsystem

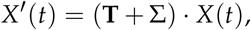

**Figure C1.**
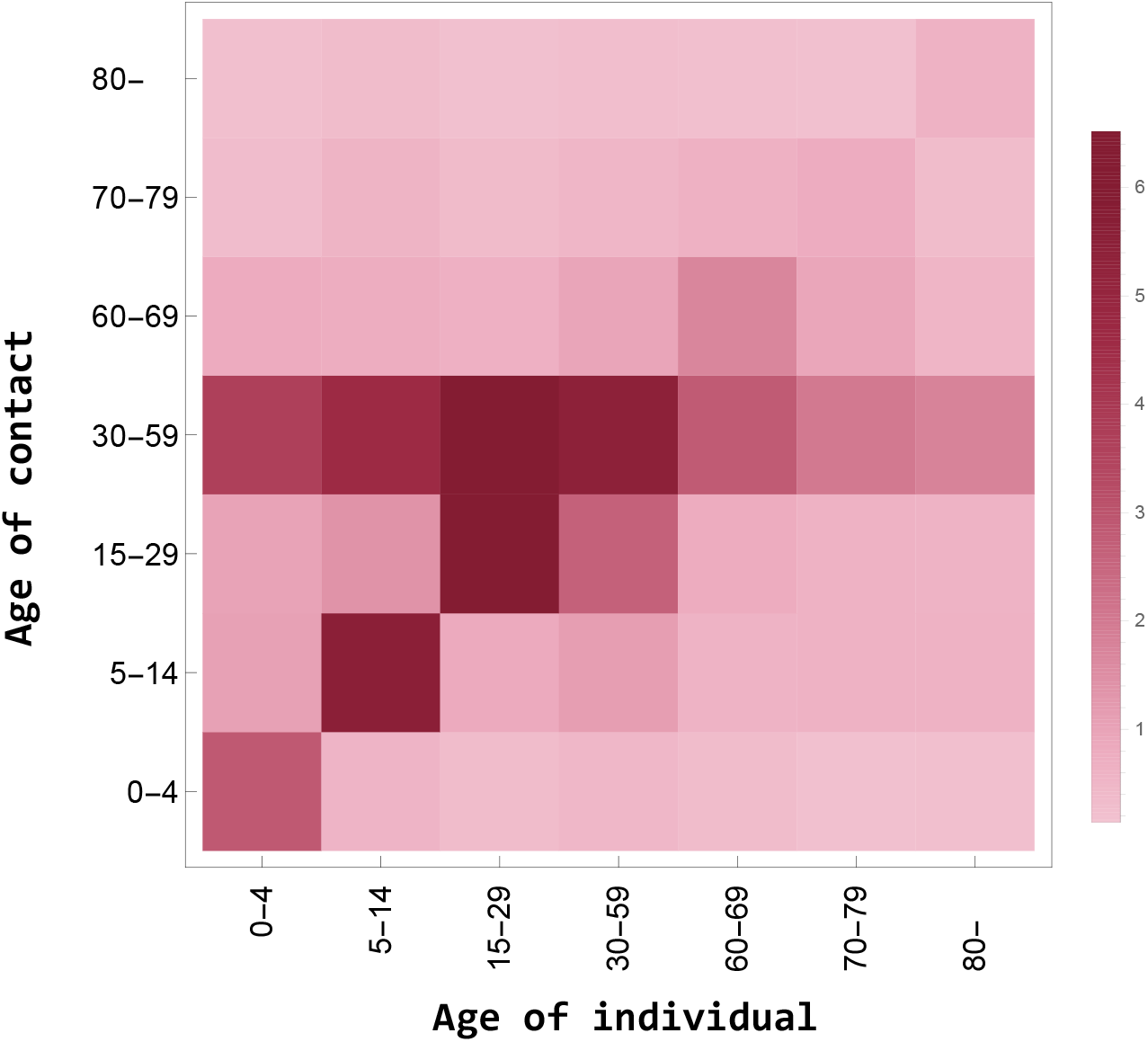
Heatmap of the contact matrix *M*_cont_

where the matrices **T** and Σ are referred to as the *transmission part* and *transitional part*, respectively; the state is described by

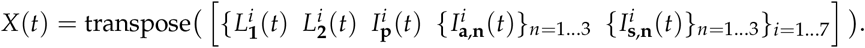

Recall that the transmission matrix **T** has the form

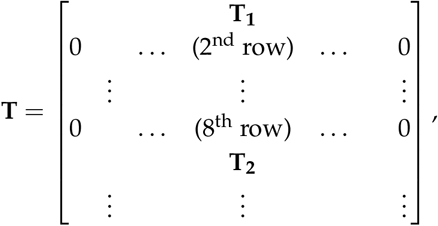

where **T_i_** = **T_1,i_ T_2,i_ T_3,i_ T_4,i_ T_5,i_ T_6,i_ T_7,i_** with

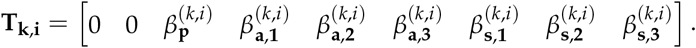

On the other hand, the transitional matrix Σ is block diagonal with blocks

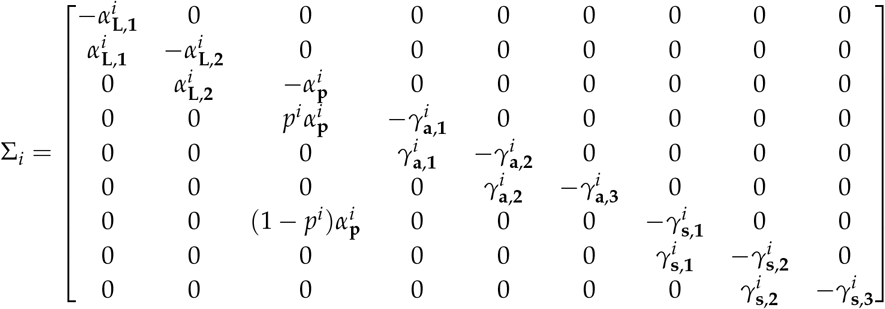

for *i* = 1,. . .,7.

Then, the NGM with large domain is given by

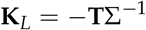

and the NGM

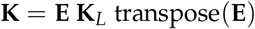

follows with the, again, block diagonal **E** with **E***_i_* =[100000000]. The baseline transmission rate *β*_0_ may be factored out from **K** as 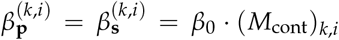 and 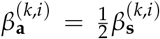. Hence, 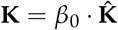, where 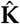may be readily constructed and we can compute its spectral radius 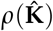. Then, we obtain the baseline transmission rate using the assumed reproduction number *R*_0_ as

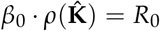

that is *β*_0_ ≈ 0.03465 for *R*_0_ = 1.65. Finally, the transmission rates 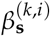 are computed via the contact matrix *M*_cont_ as 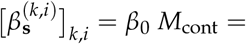

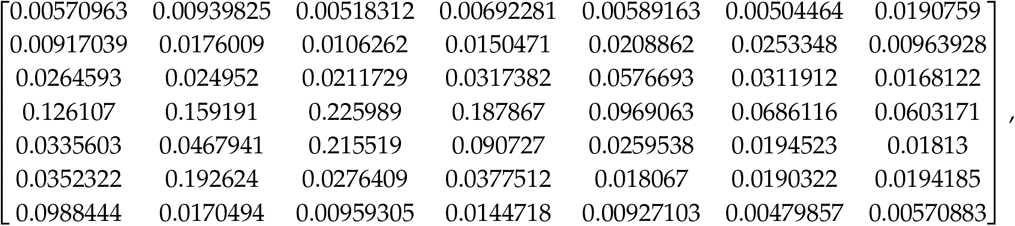

again, for *R*_0_ = 1.65. For other scenarios, the final steps are altered to align with the desired baseline reproduction number *R*_0_, resulting in an appropriate *β*_0_ and then, the scaled transmission rates 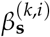.

